# Genome-wide meta-analysis for Alzheimer’s disease cerebrospinal fluid biomarkers

**DOI:** 10.1101/2022.03.08.22271043

**Authors:** Iris E Jansen, Sven J van der Lee, Duber Gomez-Fonseca, Itziar de Rojas, Maria C Dalmasso, Benjamin Grenier-Boley, Anna Zettergren, Aniket Mishra, Muhammad Ali, Victor Andrade, Céline Bellenguez, Luca Kleineidam, Fahri Küçükali, Yun Ju Sung, Niccolo Tesí, Ellen M Vromen, Douglas P Wightman, Daniel Alcolea, Montserrat Alegret, Ignacio Alvarez, Philippe Amouyel, Lavinia A Andresen, Shahram Bahrami, Henri Bailly, Olivia Belbin, Sverre Bergh, Geert Jan Biessels, Kaj Blennow, Rafael Blesa, Mercè Boada, Anne Boland, Katharina Buerger, Ángel Carracedo, Laura Cervera-Carles, Geneviève Chene, Jurgen A.H.R. Claassen, Stephanie Debette, Jean-Francois Deleuze, Peter Paul de Deyn, Janine Diehl-Schmid, Srdjan Djurovic, Oriol Dols-Icardo, Carole Dufouil, Emmanuelle Duron, Emrah Düzel, Tormod Fladby, Juan Fortea, Lutz Frölich, Pablo García-González, Maria Garcia-Martinez, Ina Giegling, Oliver Goldhardt, Timo Grimmer, Annakaisa Haapasalo, Harald Hampel, Olivier Hanon, Lucrezia Hausner, Stefanie Heilmann-Heimbach, Seppo Helisalmi, Michael T. Heneka, Isabel Hernández, Sanna-Kaisa Herukka, Henne Holstege, Jonas Jarholm, Silke Kern, Anne-Brita Knapskog, Anne M. Koivisto, Johannes Kornhuber, Teemu Kuulasmaa, Carmen Lage, Christoph Laske, Ville Leinonen, Piotr Lewczuk, Alberto Lleó, Adolfo López de Munain, Sara Lopez-Garcia, Wolfgang Maier, Marta Marquié, Merel O. Mol, Laura Montrreal, Fermin Moreno, Sonia Moreno-Grau, Gael Nicolas, Markus M Nöthen, Adelina Orellana, Lene Pålhaugen, Janne Papma, Florence Pasquier, Robert Perneczky, Oliver Peters, Yolande AL Pijnenburg, Julius Popp, Danielle Posthuma, Ana Pozueta, Josef Priller, Raquel Puerta, Inés Quintela, Inez Ramakers, Eloy Rodriguez-Rodriguez, Dan Rujescu, Ingvild Saltvedt, Pascual Sanchez-Juan, Philip Scheltens, Norbert Scherbaum, Matthias Schmid, Anja Schneider, Geir Selbæk, Per Selnes, Alexey Shadrin, Ingmar Skoog, Hilkka Soininen, Lluís Tárraga, Stefan Teipel, Betty Tijms, Magda Tsolaki, Christine Van Broeckhoven, Jasper Van Dongen, John C. van Swieten, Rik Vandenberghe, Jean-Sébastien Vidal, Jonathan Vogelgsang, Margda Waern, Michael Wagner, Jens Wiltfang, Mandy MJ Wittens, Henrik Zetterberg, Miren Zulaica, Gra@ce, EADB, Cornelia M. van Duijn, Maria Bjerke, Sebastiaan Engelborghs, Frank Jessen, Charlotte E Teunissen, Pau Pastor, Mikko Hiltunen, Martin Ingelsson, Ole Andreassen, Jordi Clarimón, Kristel Sleegers, Agustín Ruiz, Alfredo Ramirez, Carlos Cruchaga, Jean-Charles Lambert, Wiesje M van der Flier

## Abstract

Amyloid-beta 42 (Aβ42) and phosphorylated tau (pTau) levels in cerebrospinal fluid (CSF) reflect core features of the pathogenesis of Alzheimer’s disease (AD) more directly than clinical diagnosis. Initiated by the European Alzheimer & Dementia Biobank (EADB), the largest collaborative effort on genetics underlying CSF biomarkers was established, including 31 cohorts with a total of 13,116 individuals (discovery *n* = 8,074; replication *n* = 5,042 individuals). Besides the *APOE* locus, novel associations with two other well-established AD risk loci were observed; *CR1* was shown a locus for amyloid beta 42 (Aβ42) and *BIN1* for phosphorylated Tau (pTau). *GMNC* and *C16orf95* were further identified as loci for pTau, of which the latter is novel. Clustering methods exploring the influence of all known AD risk loci on the CSF protein levels, revealed 4 biological categories (amyloid, astrocyte, processing & migration, and migration & motility) suggesting multiple Aβ42 and pTau related biological pathways involved in the etiology of AD. In functional follow-up analyses, *GMNC* and *C16orf95* both associated with lateral ventricular volume, implying an overlap in genetic etiology for tau levels and brain ventricular volume.

## Introduction

Resolving the genetic background of Alzheimer’s disease (AD) has proven to contribute greatly to our understanding of underlying disease processes, for instance with the discovery of *APP* [1], *PSEN1* [2], and *PSEN2* [3] in family-based studies, leading to the amyloid cascade theory [4]. In addition, genome-wide association studies (GWAS) in AD have convincingly highlighted the importance of microglia [5, 6], a finding previously supported by research from other scientific fields [7-9], and now also widely accepted as a genetic cause rather than a result of AD pathogenesis. Further exploration of genetic risk factors contributing to AD development and pathogenesis might reveal more biological insights, an important step in the quest for AD treatment that will slow down or even halt disease progression.

GWAS of clinically diagnosed AD patients have been successful, and current efforts largely focus on increasing sample size to improve the statistical power to detect genetic variants [10, 11]. An alternative approach is to study effects of genetic variants on pathophysiological features of AD. The strength of such studies is based on the assumption that more objective measurable biological properties are more strongly associated to the underlying AD pathology than the clinical diagnostic classifications (e.g. misclassifications or symptoms not manifested yet), thereby allowing to detect larger effects by reducing heterogeneity [12]. The use of biomarkers further enables to identify genetic effects specific for certain AD-related biological mechanisms. This is an advantage over the conventional GWAS approach for clinical AD diagnosis, where it generally remains unclear through what causal gene or cellular process a locus is associated to AD.

It is possible to measure levels of amyloid-beta-42 (Aβ42) and (phosphorylated) tau (pTau and Tau) in cerebrospinal fluid (CSF), the two major proteins implicated in the AD pathological process. Aβ42 pathology in the brain is negatively correlated with CSF Aβ42 levels, where a decrease in CSF Aβ42 is indicative of AD [13, 14]. CSF (p)Tau is positively correlated with (p)Tau pathology in the brain, and therefore higher CSF (p)Tau levels are observed in patients with AD. CSF pTau is presumed to reflect AD-type tau-tangles more specifically than total tau [13, 14]. Previous studies on CSF amyloid beta and (p)Tau have identified genetic risk loci, the most recent one including 3,146 individuals [15]. Some of the 8 discovered loci had not been previously associated to AD, emphasizing the potential of endophenotypes to reveal novel genetic risk factors. Our current study aimed to further define the genetic background of AD by studying the genetic effects on CSF Aβ42 and pTau levels in a total of 13,116 individuals.

## Materials and methods

### Participants

We combined data from 16 European cohorts, encompassing a total of 8,074 individuals (Table 1, Supplementary Table 1, Supplementary Figure 1) with both genotype data and CSF measurements. The majority of these cohorts (82%) are part of the EADB consortium [10], and included the full spectrum of clinical severity potentially leading to AD, from subjective cognitive decline, mild cognitive impairment, to dementia. Written informed consent was obtained from study participants or, for those with substantial cognitive impairment, from a caregiver, legal guardian, or other proxy. Study protocols for all cohorts were reviewed and approved by the appropriate institutional review boards.

**Table 1.**
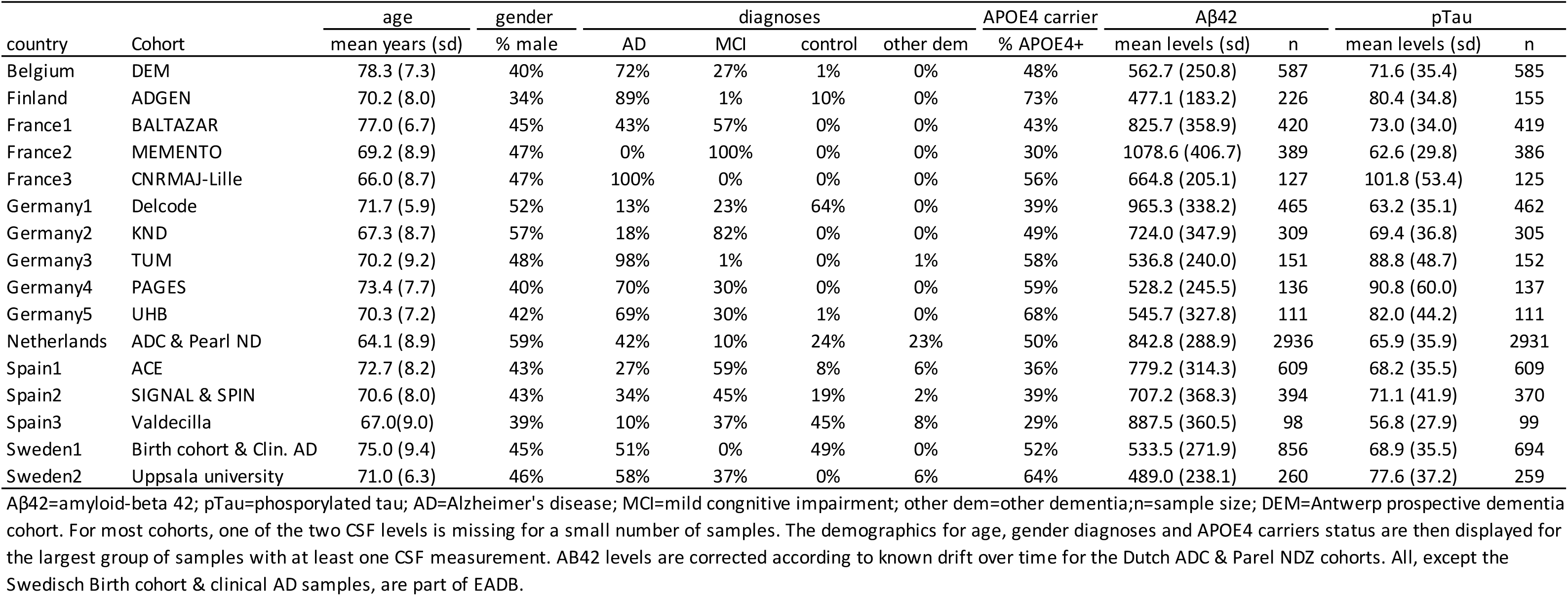
Demographic information on cohorts of stage 1 discovery analysis.

For replication, 15 cohorts totaling 5,042 individuals (Supplementary Table 2) were available to attempt replication of the association signals to Aβ42 and pTau, for the variants with p-value < 1e-5 in the discovery analysis. Data from all cohorts, except one (NorCog from University of Oslo, Norway), were obtained through collaboration with the previous largest GWAS on CSF Aβ42 and pTau, mostly including cohorts originating from the United States [15]. Basic demographics are described in Supplementary Table 2, more detailed cohort information is described elsewhere [15].

### CSF measurements

Due to the multi-center approach, CSF protein levels were measured with various CSF protein assays (Supplementary Table 2). Aβ42 was measured with ELISA, Lumipulse or V-PLEX, and pTau with ELISA or Lumipulse. For details on specific lab procedures, see the original studies [15-29]. Protein levels were log10 transformed and normalized within cohorts and CSF assay type (if multiple assays were used within a single cohort) to approximate a normal distribution to correct for the application of various CSF assays across different studies. Then, the normalized protein levels were used as continuous phenotypes in the association analyses. For the stratified analyses, two subgroups of individuals, amyloid normal and amyloid abnormal, were defined based on their Aβ42 status. Individuals with an untransformed Aβ42 level below a threshold were assigned to the abnormal amyloid level group. The thresholds were defined by the individual research groups as it depends on technical circumstances, and are displayed in Supplementary Table 2.

### Genotyping, quality control and imputation

The genetic data for the EADB cohorts has been processed in a homogeneous approach (Supplementary Table 1), in which the Illumina Infinium Global Screening Array (GSA, GSAsharedCUSTOM_24+v1.0) was predominantly used for data generation. Additional arrays included the Axiom 815K Spanish biobank array (Thermo Fisher) for ACE (Barcelona, Spain) and Valdecilla (Santander, Spain) cohorts, and the Illumina Neurochip array (Gothenburg, Sweden). Standard quality control (QC) procedures were performed to exclude individuals and variants with low quality, in general followed by imputation with the Trans-Omics for Precision Medicine (TOPMed) reference panel [30, 31]. For the EADB cohorts for which GSA genotype level data was available, the details on QC steps and imputation with the TOPMed reference panel were previously described [10].

For the Spanish ACE and Valdecilla cohorts, QC procedures are described in another study [32], followed by imputation with the TOPMed reference panel. For the Gothenburg H70 Birth Cohort studies and clinical AD samples from Sweden, the QC and imputation procedures were described elsewhere [33]. Post-imputation QC only included variants with a high imputation quality (RSQ [imputation quality] >0.8). The UCSC LiftOver program (https://genome-store.ucsc.edu/) and Plink v2.0 (www.cog-genomics.org/plink/2.0/) [34] were used to lift the GRCh37 genomic positions to GRCh38, the genomic build for all other datasets. All genotypes were hard called using the default Plink v2.0 (www.cog-genomics.org/plink/2.0/) settings.

### Heritability and genetic correlation

For the estimation of the SNP-heritability, two distinct tools were used. With LD score regression (LDSC) it was possible to perform the calculations with the full number of samples as the input for this analysis are the summary statistics. Besides heritability estimates, genetic correlations were also calculated for Aβ42, pTau, tau (to test the similarity in genetic background to pTau), and two previously published AD summary statistics [5, 35]. Precalculated LD scores from the 1,000 Genomes European reference population were obtained online. All estimates were based on HapMap3 SNPs only to ensure high-quality LD score calculations (https://alkesgroup.broadinstitute.org/LDSCORE/). As a rule of thumb, LD Score regression tends to yield very noisy results when applied to datasets with fewer than 5000 individuals (https://github.com/bulik/ldsc/wiki/FAQ) [36]. The summary statistics for the stratified analyses were therefore not considered.

For comparison to SNP-heritability estimates of previous studies for Aβ42 and pTau, GCTA v1.9 [37] was applied to the individual-level genotype data of the largest dataset (Netherlands). Other datasets were not considered as the sample size was too low for small standard errors, thereby impossible to draw any meaningful conclusions from the estimates. The restricted maximum likelihood (REML) analysis was performed for the log10-transformed normalized CSF Aβ42 and pTau adjusted for gender, age, and the first 10 principal components. Variance explained could not be calculated for significant loci only as p-values from GWAS results of a large independent sample are unavailable, and calculation in the Amsterdam sample would be hampered by winners-curse, causing inflation.

### Single-marker association analysis

Genome-wide association analysis for each cohort was performed in PLINK v2.0 [34], using linear regression for the continuous phenotypes Aβ42, tau and pTau. Association tests were adjusted for gender, age, assay type (if applicable), and ten ancestry principal components. Only variants with a minor allele frequency threshold above 0.01 were tested. For smaller cohorts (n < 250 individuals) this threshold was set to 0.05 to avoid false positive findings.

Association analyses were repeated for subgroups, stratified according to *APOE*4 status (based on the high-quality (R2>0.8) imputed variants rs429358 and rs7412) or dichotomous Aβ42 status, resulting in the following groups: 1) *APOE*4 (hetero- and homozygous) carriers; 2) *APOE*4 non-carriers; 3) individuals with abnormal Aβ42 levels; and 4) individuals with normal Aβ42 levels. After stratification, cohorts with a minimal sample size of 100 individuals were included. Covariates were those described for the main analyses above.

### Independent replication analysis

A total of 5,042 samples from 15 cohorts were included for the replication analysis. The genetic data for NorCog were generated with 2 different genotyping assays. Extensive QC procedures which are detailed elsewhere [10], allowed for joined genetic analyses of these sub-datasets. Variant association testing was performed according to the association analysis section above. For all other replication cohorts, QC, imputation and association testing procedures are described elsewhere [15]. In short, individual and variant QC standards were met, and imputation was performed using the 1000 Genomes Project Phase 3 reference panel. Each dataset was QCed and imputed independently. The additive linear regression model in PLINK v1.9 [34] was used for single-variant analyses.

### Meta-analyses

METAL [38] was used for meta-analyses in stage 1 to 3 of the per cohort association results, applying the default approach that utilizes p-value and direction of effect, while weighted according to sample size. For stage 1, we used the genome-wide threshold for significance of P<5 × 10^−8^, and a suggestive threshold of P<1× 10^−5^ to select variants to study in Stage 2. Stage 2 variants were considered a replication with P<0.05 and same direction of effect in comparison to stage 1. The genome-wide threshold for significance of P<5 × 10^−8^ was used to defined GWAS hits in stage 3.

### Colocalization

All variants within 1.5 megabases (Mb) of the lead variant of each genomic risk loci were used in the colocalization analysis. The GWAS data and eQTL data were trimmed so that all variants overlap. Colocalization was performed per gene using coloc.abf from the Coloc R package [39]. Default priors were used for prior probability of association with the GWAS data and eQTL data. The prior probability of colocalization was set as 1 × 10^−6^ as recommended [40]. Nominal P, sample size and MAF from the GWAS data and eQTL data were used in all the colocalization analyses. Colocalizations with a posterior probability >0.8 were considered successful colocalizations. eQTL data from all tissues except microglia were obtained from the eQTL catalog [41]. The microglia data were obtained from Young et al. [42]. GWAS summary statistics for loci comparison to other GWAS studies were obtained from Kunkle et al. [35] for AD, and from Vojinovic et al. [43] for brain ventricular volume.

### Gene-based analysis

Gene-based and gene-set association tests were performed using MAGMA v1.08 [44], which was implemented by FUMA [45]. The per variant association summary statistics for the main results served as the input, where variants were selected if mapped within 18,870 protein coding genes (with unique ensembl ID). The mean SNP-wise model was implemented. The Bonferroni-corrected significance threshold was set to p < 2.65×10^−6^, based on the number of tested genes.

### Gene mapping

The genome-wide significant loci of the main results were further explored for promising causal AD genes using FUMA [45], after lifting over the results with genomic build GRCh38 to GRCh37 with the UCSC LiftOver Program (https://genome-store.ucsc.edu/). Two gene mapping strategies were used:

- Positional mapping maps SNPs to genes based on physical distance (within a 10-kb window) from known protein-coding genes in the human reference assembly (GRCh37/hg19).
- eQTL mapping maps SNPs to genes with which they show a significant eQTL association (that is, allelic variation at the SNP is associated with the expression level of that gene). eQTL mapping uses information from 85 brain- and immune-related tissue types in 11 data repositories (BIOSQTL, BloodeQTL, BRAINEAC, CMC, DICE, eQTLcatalogue, eQTLGen, GTEx, PsychENCODE, scRNA_eQTLs, xQTLServer), and is based on cis-eQTLs which can map SNPs to genes up to 1 Mb apart. We used a false discovery rate of 0.05 to define significant eQTL associations.

### Phenome-wide association studies (PheWAS)

We conducted phenome-wide association studies (PheWAS) on the top SNPs, rs4844610 rs429358, rs744373, rs9877502, rs4843559. A PheWAS starts out with a single to a few variants of interest that are systematically being tested for association to many phenotypes. We used the ‘phewas’ function of the R-package ‘ieugwasr’ [46, 47]. Using this function, we searched traits that associate with the list of SNPs with P<1×10-7 in all GWAS harmonized summary statistics in the MRC IEU OpenGWAS data infrastructure [47]. In short, this enables us to screen for other traits to which these SNPs are associated. The database (May 2021) includes the GWAS summary statistics of 19,649 traits.

### Association with CSF proteomics

We associated the lead variants near *GMNC* (rs9877502) and in *C16orf95* (rs4843559) with CSF proteomics data of two different sources (EMIF-AD MBD and Knight-ADRC). For the EMIF-AD MBD data, a total of 2,136 proteins were quantified centrally using 11-plex tandem mass tag spectrometry in 366 individuals from the EMIF-AD MBD study [48] (subset of Amsterdam Dementia Cohort within EADB). We selected proteins with a maximum of 50% missing values. For related proteins that had identical values due to fragment aspecificity, we randomly selected one protein for analysis (52 proteins were excluded). Out of the 2,136 proteins quantified, 1,282 (55.4%) proteins respected these criteria and were included in the study.

For the Knight-ADRC data, levels of 1,305 proteins were quantified using the SOMAscan assay, a multiplexed, aptamer-based platform CSF (n = 717) [49]. Quality control was performed at the sample and aptamer levels using control aptamers (positive and negative controls) and calibrator samples. As described in detail [49], additional quality control was performed that included limit of detection cut-off, scale factor, coefficient of variation, and outlier variation. Only proteins with a call rate higher than 85% call rate were included. A total of 713 proteins passed quality control. pQTL analyses was performed and reported in previous studies [49].

g:Profiler and Enrichment map [50], a Cytoscape App, were used to perform pathway enrichment analyses on proteins with a certain level of association (EMIF-AD MBD: P<0.05; Knight-ADRC: P< 0.004, corresponding to proteins with a similar effect size as in EMIF-AD MBD). The results are shown as functionally grouped networks. We used GO biological processes and Reactome as ontology sources. For this explorative analysis, only pathways with P<0.05 (corrected for multiple testing) are shown.

### Effects of AD-associated variants on Aβ42 and pTau

We assessed the most recent GWAS [10] for AD and extracted the top loci of 83 variants (excluding *APOE* ε4 and *APOE* ε2) that showed genome-wide significant association with AD [10]. We extracted Z-scores and p-values and plotted them in a heatmap. Rows and columns were clustered using Euclidean distances and average hierarchical clustering. We performed a gene-set enrichment analysis to find molecular pathways enriched within each cluster. The SNP-gene assignment corresponds to the one described in the recent main EADB GWAS [10], including several annotation strategies. When multiple genes were reported to associate with the same SNP (rs12590654 near *SLC24A4*/*RIN3*, rs7225151 near *SCIMP*/*RABEP1*, rs6846529 near *CLNK*/*HS3ST1*, rs7384878 near *ZCWPW1*/*NYAP1* and rs10437655 near *CELF1*/*SPI1*), we considered both genes for the gene-set enrichment analysis. In addition, for SNP rs6605556, located in the complex HLA region, we considered HLA-DRB1 gene (eQTL in blood with rs6605556), and for SNPs rs7157106 and rs10131280, both located in the gene-dense IGH region, we considered IGHG2 and IGHV2-70 (eQTLs in blood with rs7157106 and rs10131280, respectively). The gene-set enrichment analysis was performed specifying Biological Processes from Gene Ontology [51, 52] as gene-set and correcting p-values with Bonferroni. Biological pathways were considered significant at corrected p<0.05. To help with the interpretation of each cluster’s function, we plot the most recurring words of the significant terms underlying each cluster using wordclouds. The following R packages were used for these analysis: gprofiler2 [53] and wordcloud2 (https://github.com/lchiffon/wordcloud2).

## Results

The overview of the study design is illustrated in Supplementary Figure 1. The GWA results from 16 studies were combined in stage 1. Variants that reached a suggestive level of significance (p<1×10^−5^) were subsequently evaluated in an independent sample from 15 studies in stage 2. Finally, the results of stage 1 and stage 2 analyses were combined in stage 3. Detailed information on study participants, CSF acquisition and genotyping is provided in Table 1 and Supplementary Table 1 and 2. The results for tau and pTau are strongly correlated (r_g_=0.94; *p*=1.86×10^−118^), and therefore only pTau findings are reported.

### Genetic architecture

The fraction of variance in Aβ42 and pTau protein levels that could be explained by the additive effect of the genetic variants tested, was estimated on 0.13 (SE=0.06) and 0.21 (SE=0.07) by LDSC, respectively. These SNP-heritabilities are substantially higher than the 0.07 previously estimated with LDSC for the diagnosis AD [54], or similarly reported for AD by this study using the same LDSC method on more recent public GWAS summary statistics of AD [5, 35] (Supplementary Table 3). GCTA estimated the SNP-heritability to be higher for both Aβ42 and pTau, namely 0.27 (SE=0.13) and 0.34 (SE=0.12), respectively. Both methods are reporting a higher SNP-heritability for pTau than for Aβ42. Genetic correlation estimates with AD GWAS summary statistics are described in the Supplementary Results and Supplementary Table 4.

### GWAS variants associated with CSF Aβ42 and pTau

The stage 1 meta-analyses (QQ plots and lambda shown in Supplementary Figure 2) identified 4 independent significant variant associations, 1 for Aβ42 and 3 for pTau (Table 2). The strongest associations were observed for both Aβ42 and pTau in the *APOE* locus (Table 2). The variant that determines the *APOE* ε4 allele (rs429358-C) decreased Aβ42 (Z=-36.29; *p* = 2.0×10^−288^) and increased pTau (Z=18.31; *p*=6.87×10^−75^) in CSF. In contrast, the variant determining the *APOE* ε2 haplotype (rs7412-T) increased Aβ42 (Z=11.97; *p*=5.09×10^−33^) and decreased pTau (Z=-6.59; *p*=4.49×10^−11^). The *APOE* ε2 association was replicated in stage 2 (Aβ42: Z=7.27; *p*=3.73×10^−13^, and pTau: Z=-6.43; *p*=1.26×10^−10^), and *APOE* ε4 with rs4420638-G for Aβ42 (Z=-25.51; *p*=1.57×10^−143^), and with rs769449-A for pTau (Z=13.83; *p*=1.66×10^−43^), both variants in high linkage disequilibrium with rs429358, as the original *APOE* ε4 variant was not genotyped or imputed in the replication datasets.

**Table 2.**
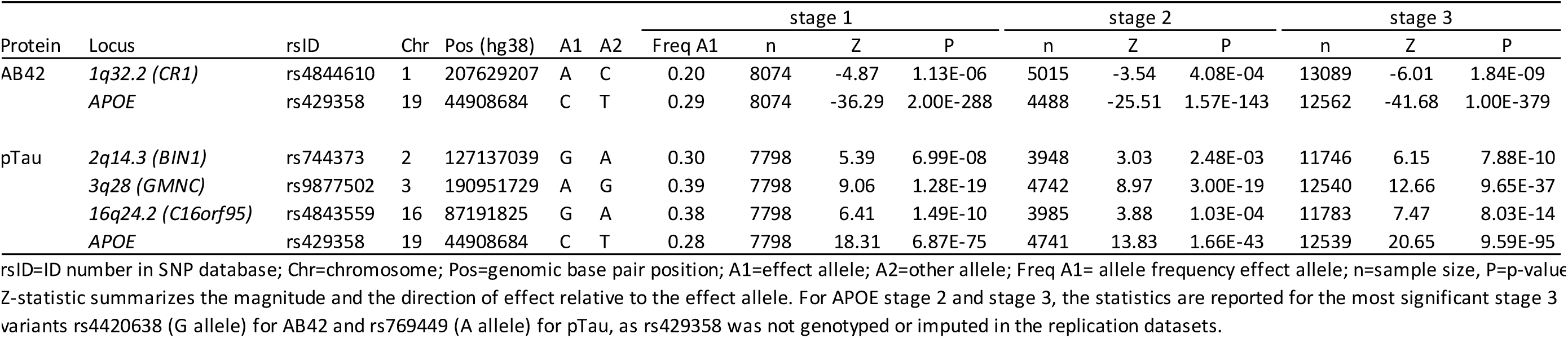
Meta-analysis association results for the 3 stages.

In stage 1, no other significant loci were observed for Aβ42. For pTau, we further identified significant associations mapping to two chromosomal regions at 3q28 and 16q24.2 (Figure 2, Table 2). The 3q28 locus (Z=9.06; *p*=1.28×10^−19^) also known as the *GMNC* locus, was reported previously for its association with pTau [15]. The cohorts from this previous study are the replication cohorts of the current study, thereby logically *GMNC* was replicated (Z=8.97; *p*=3.00×10^−19^). The 16q24.2 locus (Z=6.41; *p*=1.49×10^−10^), which is novel for pTau, was replicated in the stage 2 meta-analysis (Z=3.88; *p*=1.03×10^−04^).

Subsequently, the results from all individual studies were combined in the stage 3 meta-analysis (N =13,116). In stage 3, two well-known AD loci showed additional genome-wide significant associations (Figure 1, Table 2) with Aβ42 in chromosomal region 1q32.2 (Z=-6.01; *p*=1.84×10^−9^, *CR1*), and with pTau for the region 2q14.3 (Z=6.15; *p*=7.88×10^−10^, *BIN1*). The per-cohort and zoomed in genomic location details of all significantly associated loci of stage 3 are visualized in Supplementary Figures 3 to 8. Colocalization analyses (results detailed in Supplementary Table 5) showed colocalization (posterior probability = 0.85) of the *CR1-*locus to the *CR1-*locus of a recent AD GWAS [35]. The *BIN1-*locus did not colocalize (posterior probability = 0.01), thereby suggesting that other causal variants are underlying the association signal (posterior probability = 0.99).

**Figure 1.**
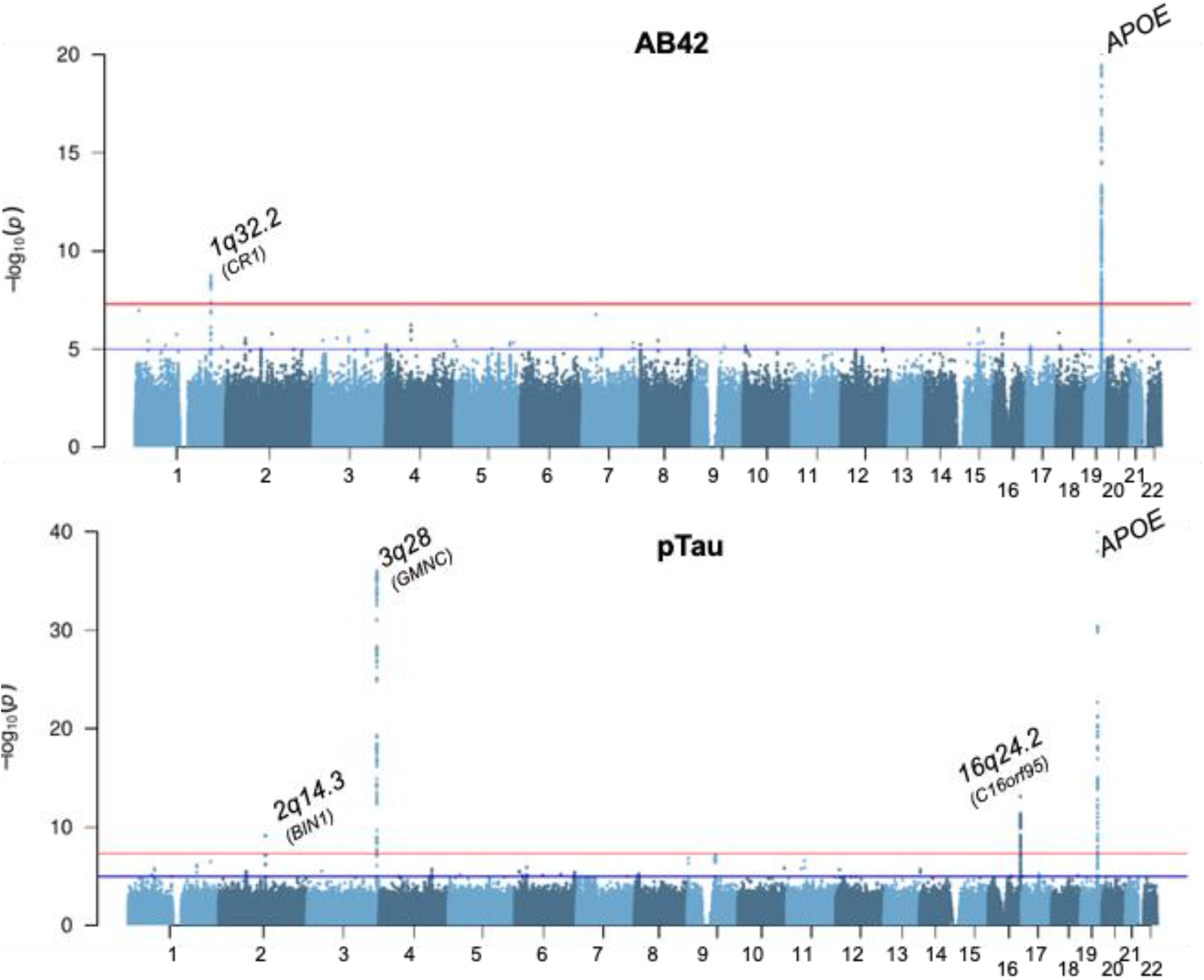
Manhattan plots of the stage 3 GWAs for Aβ42 and pTau. The y-axes are limited to visualize the non-APOE loci. The lowest p-values for APOE are 4.07×10^−355^ and 3.74×10^−94^ for Aβ42 and pTau, respectively.

Explorative meta-analyses were repeated stratified for *APOE* (*APOE* ε4 carriers (n=3,240) vs. *APOE* ε4 non-carriers (n=3,201)) and amyloid status (Amyloid normal levels (n=3,182) vs. amyloid abnormal levels (n=3,775)) for stage 1 (QQ plots and lambda shown in Supplementary Figure 9 and 10), of which the results are visualized in Supplementary Figures 11 and 12, and detailed in Supplementary Results and Supplementary Table 6. Besides the *APOE* and *GMNC* loci, two novel loci are observed that have previously not been linked to any AD phenotype.

### Functional interpretation

To interpret the functional effects of the identified variants beyond AD, we performed gene prioritization (based on positional mapping, gene-based association results, and brain and immune eQTL annotations) using FUMA [45], colocalization analyses and PheWAS. The results of the FUMA annotation are detailed in the Supplementary Results and Supplementary Table 7. None of our CSF Aβ42 and pTau loci colocalized with any of the brain or immune eQTLs from the 41 tested datasets.

For the PheWAS, using data from publicly available genome-wide association studies (N= 19,649) of the five top variants yielded 529 associations at p<1×10-7 (Supplementary Table 8). The majority is the known wide range of 490 traits associations with the APOE ε4 allele. For the other variants 39 associations were reported for 27 unique traits. These traits can be categorized in three groups: traits related to brain ventricular volumes in particular the lateral-ventricle (*GMNC* and *C16orf95*), Alzheimer’s disease diagnosis (*BIN1* and *CR1*), and measures of blood cell/lymphocyte counts (*CR1*). The regional pTau associations of *GMNC* and *C16orf95* overlapped with ventricular volume (Figure 2). Colocalization results (Supplementary Table 5) implies the same causal variant for *C16orf95* (posterior probability=0.85), though based on the current analysis it seems the *GMNC* locus cannot be explained by the same causal variant (posterior probability=0.25). This suggests that different causal variants are underlying the *GMNC* association signals for both phenotypes, or that multiple causal variants are at play, which is currently not tested in the colocalization model (as it assumes a single causal variant).

**Figure 2.**
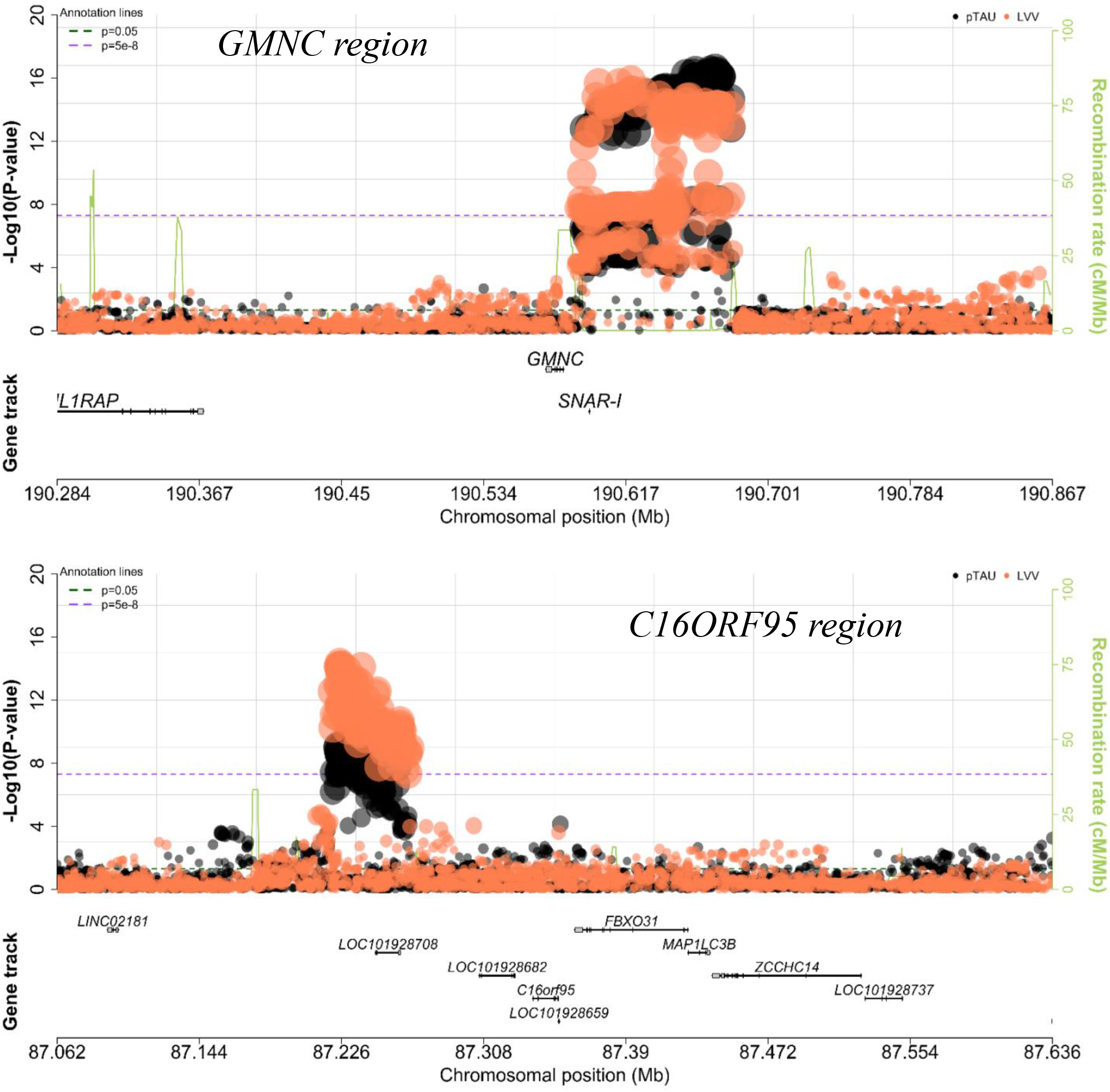
LocusZoom plots showing variant association results for *GMNC* and *C16orf95* loci. In black the pTau association signals of this study, and in orange the lateral ventricular volume (LVV) association signals observed in other studies.

Because of the overlap in effect of *GMNC* and *C16orf95* we hypothesized that these two loci affect the same biological pathways. We explored this hypothesis using CSF proteomics datasets of the EMIF consortium with 1,284 quantified proteins and of Knight-ADRC with 696 quantified proteins (of which 42% overlap with the EMIF-AD MBD proteins). For *GMNC* there were 279 (22%) proteins associated in the EMIF-AD MBD data (Supplementary Table 9) and 255 (36%) proteins in the Knight-ADRC data (Supplementary Table 10). *C16orf95* could only be tested in the EMIF-AD MBD data in which 73 (6%) proteins were associated (Supplementary Table 9). Only 2 proteins (CDH9 and DPP6) overlapped between the 2 loci. We studied the overlap in affected pathways between the associated protein lists. For *GMNC*, consistent functional group networks between the 2 tested datasets were axon guidance and ephrin signaling (Supplementary Figure 13, Supplementary Tables 11 and 12), while for *C16orf95* (only based on the EMIF-AD MBD data) glycosaminoglycan metabolism and ECM organization were overrepresented functional groups (Supplementary Figure 14, Supplementary Table 13). There was little overlap between the loci in the pathways that emerged from the protein lists.

### Relation to AD associated genetic variants

Because of the evident overlap in etiology with clinical AD dementia we explored the association of all known AD loci (excluding the APOE locus) with CSF Aβ42 and pTau. The results are shown in the heatmap of Figure 3 and Supplementary Tables 14 and 15. The variants could be clustered in 4 groups of AD-associated genes based on their associations with Aβ42 and pTau. The first cluster of 14 variants showed strong association with both decreased levels of Aβ42 and increased levels of pTau in CSF. A pathway enrichment analysis of the genes associated with the variants showed 19 GO terms enriched and ‘amyloid’ is the common denominator in the names of these terms. The second cluster contained 21 variants and included genes that have also been related to other dementia types (e.g. *GRN, TMEM107B, SNX1, MAPT, CTSB* and *CTSH*). This cluster was associated with decreased pTau levels, an no general effect on Aβ42 levels. Pathway analysis of the genes suggests an enrichment for 4 GO terms of which the names have ‘astrocyte’ as a common denominator. The third cluster consisted of 22 genes, which were related to decreased levels of Aβ42 but not increased levels of pTau. Seven GO-terms were enriched and ‘processing’ and ‘migration’ are the words that occur most often in these terms. The last cluster of 20 genes group because they increased pTau, but did not decrease Aβ42 levels. Ten GO terms are significantly enriched in this gene cluster and ‘migration’ and ‘motility’ are most common words in these GO-terms.

**Figure 3.**
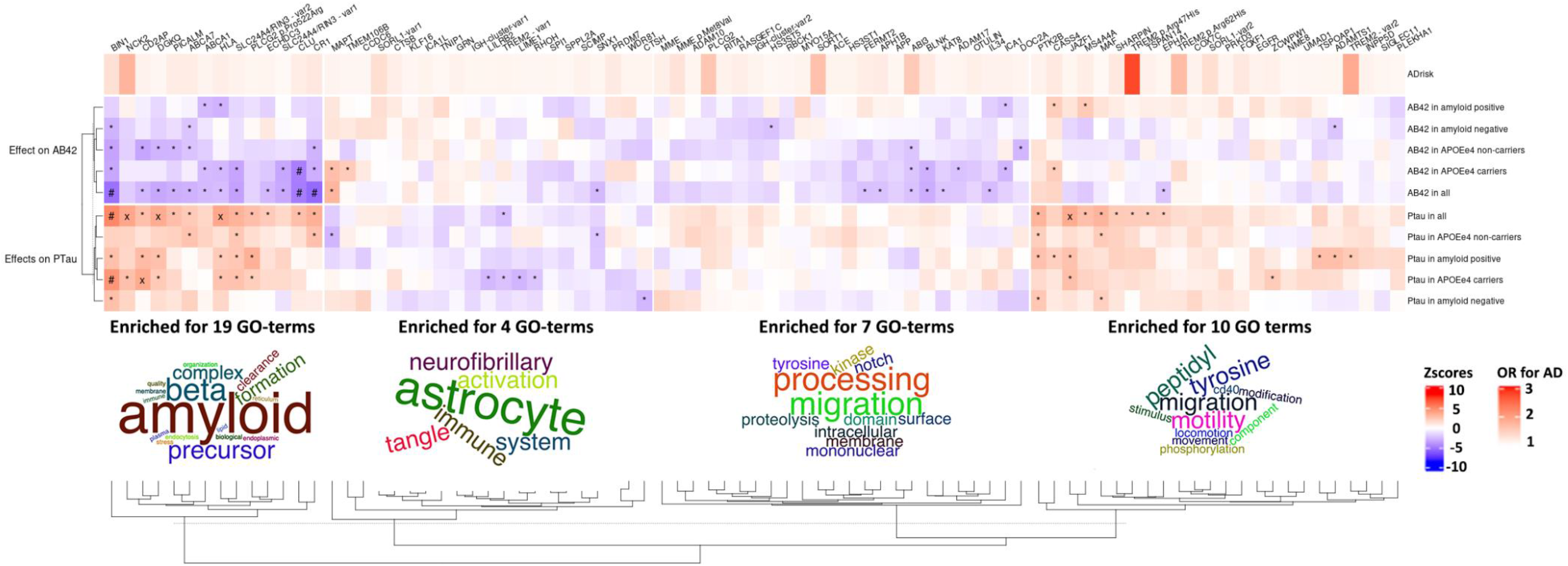
The effects of all AD associated loci. The names of the loci are named according to their linked gene names in Bellinguez et al. (2021). Hierarchical clustering was performed on the rows and columns using Eucledian distances and the method ‘average’ for clustering Pathway enrichment analyses were performed on the four first clusters. The enrichment analyses are in Supplementary Table 9. The upper bar shows the odds ratio for AD, where alleles for variants with protective effects have been flipped to show AD-risk increasing effects for all variants. The increases in Aβ42 and pTau (positive Zscores) are shown in red and decreases in AB42 and PTau (negative Zscores) are shown in blue. * = p-value <0.05, X = 0.05 < p-value <0.001, # = 0.001 < p-value <5×10-8, $ = p-value <5×10-8.

## Discussion

We identified 2 loci (*CR1* and *APOE*) for Aβ42, and 4 loci (*BIN1, GMNC, C16orf95* and *APOE*) for pTau in a total of 13,116 individuals (discovery *n* = 8,074; replication *n* = 5,402 individuals). In concordance with previous studies [15, 55], both proteins showed the strongest association for *APOE*, where *APOE ε*4 decreased amyloid beta levels and increased pTau levels, while *APOE* ε2 had the opposite effect. We confirmed *GMNC* as a risk factor for CSF pTau levels. We identified *CR1* as a novel locus for CSF Aβ42 levels, and we observed 2 novel loci (*BIN1* and *C16orf95*) for CSF pTau levels. So other than *APOE*, no risk loci overlap is observed for Aβ42 and pTau, implying at least partly separate genetic backgrounds for both pathological hallmarks. Amyloid beta appears to be dominated by the effect of *APOE*, while pTau is influenced by multiple genetic components. Such a divergence in genetic influences is not in concordance with a genetic etiology where accumulation of Tau tangles is a direct downstream effect of amyloid plaque formation, as proposed by the amyloid cascade theory [56, 57]. Rather, it seems that these pathologies originate at least partly independent from each other, which is highly relevant biological knowledge for the development of potential AD treatments. The limited clinical efficacy of agents that aim to reduce beta-amyloid plaques might potentially be due to this [58]. In line with this implication is the observed difference in genetic clusters based on CSF protein patterns, suggesting multiple Aβ42 and pTau related biological pathways (related to amyloid, astrocyte, migration & processing, and migration and motility) to be involved in the etiology AD. The variety in subclasses of genetic contributors for AD etiology could mean that different patient groups might benefit from distinct AD treatment depending on the biological pathway that is affected.

Our *CR1* findings, are in agreement with the well-known observed association for AD risk, where rs6656401 (R^2^ = 0.88 with lead SNP of current study) carriers are more susceptible for AD. The similarity in these association signals is strengthened by the convincing colocalization of the *CR1* locus between our CSF pTau observation and the *CR1* locus of Kunkle et al. [35]. Functional work on the effect of CR1 further suggests that CR1 is involved in AD pathogenesis by regulating Aβ42 clearance, both peripherally with blood cells and in the brain itself [59]. More recent research in red blood cells of AD patients showed deficient CR1 immunoreactivity, including CR1-mediated capture of circulating amyloid beta [60]. They observed decreased CR1 protein levels in red blood cells for *CR1* SNPs that associate with higher AD risk. The second novel locus in this study is *BIN1* for CSF pTau. Of note is the colocalization result suggesting a different causal variant for AD and pTau, which could have important implications for functional understanding of BIN1’s functional involvement in AD via pTau. First, more in-depth comparison with different models (e.g. multiple causal variants assumption) between these two loci should be performed before valuable functional interpretation is possible. *BIN1* has already been linked to Tau pathology in several functional studies, first shown in fruit flies where a decrease in the *BIN1* ortholog gene expression suppressed Tau-mediated neurotoxicity [61]. More recently, research in mice showed physical protein interaction between BIN1 and Tau [62], and BIN1 involvement in Tau-dependent hyperexcitability in AD [63]. In human subjects, *BIN1-*carriers were associated with lower memory performance, mediated by higher tau-PET levels [64]. Our observed *BIN1-*Tau association contributes valuable knowledge on this topic, by observing the same trend in a substantial larger study (89 vs 13,118 individuals) using different techniques to measure Tau pathology (PET vs. CSF). We provide in vivo confirmation that *CR1* is associated with AD via Aβ42, while *BIN1* relates to Tau pathology.

The third novel locus in this study is the region on genomic location 16q24.2 for pTau, of which the strongest associated variants are located within intronic regions of *C16orf95*. This locus has not been linked to (p)Tau pathology or AD in previous research, though it associated to lateral ventricular volume in the CHARGE study, including 23.5k healthy individuals [43]. Similarly, the 3q28-locus for pTau from our findings has also been associated to lateral ventricular volume by the CHARGE study, implying that the same genetic risk factors contribute to both phenotypes, strengthening the notion that neurodegeneration and (p)Tau pathology are highly correlated. 3q28 has been linked to (p)Tau by previous CSF studies in dementia cohorts and was identified as the *GMNC-*locus [15, 55]. In comparison to the latest GWAS of Deming et al. [15], increasing the sample size with a small 10k individuals in this study strengthens the association of this locus from 3.07×10^− 11^ to 1.19×10^−36^, thereby turning it into a well-established locus for pTau pathology (similar for Tau: Deming *p*=3.07×10^−11^ (n=3,146); current *p*=9.65×10^−37^ (n=12,540). The formerly reported directions of effect of *GMNC* and *C16orf95* for ventricular volume are counterintuitive. For both loci the allele that associated with an increase in pTau pathology in our dementia cohorts associated with a smaller ventricular volume, implying less neurodegeneration. We explored if these loci work through the same biological pathways using CSF proteomics data. The consistently highlighted functional groups for the *GMNC* locus (axon guidance and ephrin signaling) were different than for the *C16orf95* locus (extracellular matrix components), thereby implying at least partly distinct functional routes via which they influence pTau protein levels in CSF.

We furthermore identified two novel loci in the stratified analyses for Aβ42 levels: 7q11.22 for *APOE* ε4 non-carriers and 12q13.3 in individuals with abnormal amyloid levels. We were unable to test for replication of these loci in independent replication datasets, as such analyses were not performed. Neither loci have been previously linked to AD, or any other trait. The lead SNP for the locus on chromosome 7 is a common intronic variant for the lncRNA *LOC105375341*, which according to GTEx is only expressed in testis and prostate, and thereby not a promising causal gene for AD or AD-related phenotypes. The locus on chromosome 12 consists of just one rare intronic variant that was only detected in the Dutch cohort with the largest sample size (n=498). Future studies including more cohorts of large sample sizes are required to study this rare variant in more detail.

Besides *APOE* and *GMNC*, no other loci from the latest GWAS of CSF amyloid and tau [15] were replicated by this study. For Aβ42, it was not possible to test the *GLIS1*-locus as this variant was not observed in our data, which is in concordance with the gnomAD browser[65] reporting extremely low coverage and a MAF of 0.007 for rs185031519, the strongest *GLIS1* SNP in Deming et al. For the other loci that we were unable to replicate (*SERPINB1* for Aβ42; and *GLIS3, PCDH8, CTDP1* for pTau), there were no significant associations (p>0.05) despite substantial sample sizes (n>7000). These differences in findings might be due to differences in study design, for example inclusion of cohorts with other diagnoses and/or differences in analysis strategies.

The GCTA-based SNP-heritability estimates of 27% and 34% for Aβ42 and pTau, respectively, are in a similar range to the estimates calculated by Deming et al [15] (36% for Aβ42 and 25% for pTau). Notable is that the previous study estimated Aβ42 to be most heritable, while we observed the highest estimate for pTau, though the standard errors were about 10% for both studies. Our LDSC-based SNP-heritabilities show a similar trend with higher estimates for pTau. Furthermore, these LDSC-based 13% for Aβ42 and 21% for pTau protein levels are considerably higher than the 7% previously observed for the diagnosis AD. The higher heritability for the tested CSF protein levels strengthens the assumption that more objective measurable biological properties are more strongly associated to AD pathogenesis than the diagnostic classifications.

In conclusion, the current findings clearly show that studying the genetic effects of AD-related endophenotypes has the potential to reveal novel associations, and highlight important biological insights. The clear distinction in genetic findings for amyloid-beta and tau emphasizes the (partly) genetic independence of these two biological mechanisms in AD pathogenesis. Moreover, the identification of *CR1* and *BIN1*, which are the second and third strongest associated AD locus after *APOE*, furthermore implies that by increasing sample size of genetic analysis in CSF biomarkers it will become more apparent through which biological mechanisms certain AD loci have their effect on AD pathogenesis. Even larger collaborative efforts with more homogeneous sample definitions are therefore encouraged to be undertaken to enhance our genetic understanding of AD, ultimately leading to improved biological knowledge for the development of drug treatment.

## Supporting information

Supplemental text, figures and authors.

Ethical committees table

## Data Availability

All data produced will be made available online upon acceptance of the manuscript for publication by a peer-reviewed journal.

## Acknowledgments

This work was supported by a grant (European Alzheimer DNA BioBank, EADB) from the EU Joint Programme, Neurodegenerative Disease Research (JPND). Amsterdam dementia Cohort (ADC): Research of the Alzheimer center Amsterdam is part of the neurodegeneration research program of Amsterdam Neuroscience. The Alzheimer Center Amsterdam is supported by Stichting Alzheimer Nederland and Stichting VUmc fonds. The clinical database structure was developed with funding from Stichting Dioraphte. Genotyping of the Dutch case-control samples was performed in the context of EADB (European Alzheimer DNA biobank) funded by the JPco-fuND FP-829-029 (ZonMW projectnumber 733051061).Part of the work described in this study was carried out in the context of the Parelsnoer Institute (PSI). PSI was part of and funded by the Dutch Federation of University Medical Centers and has received initial funding from the Dutch Government (from 2007-2011). Since 2020, this work was carried out in the context of Parelsnoer clinical biobanks at Health-RI (https://www.health-ri.nl/initiatives/parelsnoer). Part of the genotyping included in this work was funded by the JPND EADB grant (German Federal Ministry of Education and Research (BMBF) grant: 01ED1619A). Alfredo Ramirez is also supported by the German Research Foundation (DFG) grants Nr: RA 1971/6-1, RA1971/7-1, and RA 1971/8-1.

We would like to thank patients and controls who participated in this project. The Genome Research @ Fundació ACE project (GR@ACE) is supported by Grifols SA, Fundación bancaria ‘La Caixa’, Fundació ACE, and CIBERNED. A.R. and M.B. receive support from the European Union/EFPIA Innovative Medicines Initiative Joint undertaking ADAPTED and MOPEAD projects (grant numbers 115975 and 115985, respectively).M.B. and A.R. are also supported by national grants PI13/02434, PI16/01861, PI17/01474, PI19/01240 and PI19/01301. Acción Estratégica en Salud is integrated into the Spanish National R + D + I Plan and funded by ISCIII (Instituto de Salud Carlos III)–Subdirección General de Evaluación and the Fondo Europeo de Desarrollo Regional (FEDER–’Una manera de hacer Europa’). The position held by I.dR. is funded by grant. FI20/00215. PFIS Contratos Predoctorales de Formación en Investigación en Salud.

We would like to thank UCL Genomics, London, UK, for performing the genotyping analyses of the samples within the Gothenburg H70 Birth Cohort Studies and Clinical AD Sweden. The recruitment and clinical characterization of research participants at Washington University were supported by NIH P30AG066444, and P01AG003991. This work was supported by access to equipment made possible by the Hope Center for Neurological Disorders, the Neurogenomics and Informatics Center (NGI: https://neurogenomics.wustl.edu/)and the Departments of Neurology and Psychiatry at Washington University School of Medicine. Research at the Belgian EADB site is funded in part by the Alzheimer Research Foundation (SAO-FRA), The Research Foundation Flanders (FWO), and the University of Antwerp Research Fund. FK is supported by a BOF DOCPRO fellowship of the University of Antwerp Research Fund. The work of Valdecilla was supported by grants from the Instituto de Salud Carlos III (Fondo de Investigación Sanitario, PI08/0139, PI12/02288, PI16/01652, and PI20/01011), the JPND (DEMTEST PI11/03028), the CIBERNED, and the Siemens Healthineers. We thank the Valdecilla Biobank (PT17/0015/0019), integrated into the Spanish Biobank Network, for their support and collaboration in sample collection and management. Our heartfelt thanks to the participants of the Valdecilla Cohort for their generosity. The work of ADGEN was supported by EU Joint Programme - Neurodegenerative Disease Research (301220) and the Academy of Finland (338182). The DELCODE study (Study-ID:BN012) was supported and conducted by the German Center for Neurodegenerative Diseases (DZNE). The data samples were provided by the DELCODE study group. Details and participating sites can be found at www.dzne.de/en/research/studies/clinical-studies/delcode. The German Dementia Competence Network (KND) is funded by the German Federal Ministry of Education and Research (BMBF) grants Number: 01G10102, 01GI0420, 01GI0422, 01GI0423, 01GI0429, 01GI0431, 01GI0433, 04GI0434, 01GI0711.

WF, SvdL, HHolstege, CT and PhS are recipients of ABOARD, which is a public-private partnership receiving funding from ZonMW (#73305095007) and Health∼Holland, Topsector Life Sciences & Health (PPP-allowance; #LSHM20106). More than 30 partners participate in ABOARD (www.aboard-project.nl). ABOARD also receives funding from de Hersenstichting, Edwin Bouw Fonds and Gieskes-Strijbisfonds. IEJ was partially supported by NWO Gravitation program BRAINSCAPES: A Roadmap from Neurogenetics to Neurobiology (NWO: 024.004.012). AZ was supported by the Swedish Alzheimer Foundation (AF-939988, AF-930582, AF-646061, AF-741361), and the Dementia Foundation (2020-04-13, 2021-04-17). ISk was supported by the Swedish state under the agreement between the Swedish government and the county councils, the ALF-agreement (ALF 716681), the Swedish Research Council (no 11267, 825-2012-5041, 2013-8717, 2015-02830, 2017-00639, 2019-01096), Swedish Research Council for Health, Working Life and Welfare (no 2001-2646, 2001-2835, 2001-2849, 2003-0234, 2004-0150, 2005-0762, 2006-0020, 2008-1229, 2008-1210, 2012-1138, 2004-0145, 2006-0596, 2008-1111, 2010-0870, 2013-1202, 2013-2300, 2013-2496), Swedish Brain Power, Hjärnfonden, Sweden (FO2016-0214, FO2018-0214, FO2019-0163), the Alzheimer’s Association Zenith Award (ZEN-01-3151), the Alzheimer’s Association Stephanie B. Overstreet Scholars (IIRG-00-2159), the Alzheimer’s Association (IIRG-03-6168, IIRG-09-131338) and the Bank of Sweden Tercentenary Foundation. SK was supported by the Swedish state under the agreement between the Swedish government and the county councils, the ALF-agreement (ALFGBG-81392, ALF GBG-771071), the Swedish Alzheimer Foundation (AF-842471, AF-737641, AF-939825), and the Swedish Research Council (2019-02075). MW was supported by the Swedish Research Council 2016-01590. HZ is a Wallenberg Scholar supported by grants from the Swedish Research Council (2018-02532), the European Research Council (681712), Swedish State Support for Clinical Research (ALFGBG-720931), the Alzheimer Drug Discovery Foundation (ADDF), USA (201809-2016862), the European Union’s Horizon 2020 research and innovation programme under the Marie Skłodowska-Curie grant agreement No 860197 (MIRIADE), and the UK Dementia Research Institute at UCL. KB was supported by the Swedish Research Council (#2017-00915), the Alzheimer Drug Discovery Foundation (ADDF), USA (#RDAPB-201809-2016615), the Swedish Alzheimer Foundation (#AF-742881), Hjärnfonden, Sweden (#FO2017-0243), the Swedish state under the agreement between the Swedish government and the County Councils, the ALF-agreement (#ALFGBG-715986), the European Union Joint Program for Neurodegenerative Disorders (JPND2019-466-236), the National Institute of Health (NIH), USA, (grant #1R01AG068398-01), and the Alzheimer’s Association 2021 Zenith Award (ZEN-21-848495). CC receives support from the National Institutes of Health (R01AG044546, R01AG064877, RF1AG053303, R01AG058501, U01AG058922, RF1AG058501, R01AG064614), and the Chuck Zuckerberg Initiative (CZI).

## Competing Interests

Research programs of Wiesje van der Flier have been funded by ZonMW, NWO, EU-FP7, EU-JPND, Alzheimer Nederland, CardioVascular Onderzoek Nederland, Health∼Holland, Topsector Life Sciences & Health, stichting Dioraphte, Gieskes-Strijbis fonds, stichting Equilibrio, Pasman stichting, stichting Alzheimer & Neuropsychiatrie Foundation, Biogen MA Inc, Boehringer Ingelheim, Life-MI, AVID, Roche BV, Fujifilm, Combinostics. WF holds the Pasman chair. WF is recipient of ABOARD, which is a public-private partnership receiving funding from ZonMW (#73305095007) and Health∼Holland, Topsector Life Sciences & Health (PPP-allowance; #LSHM20106). WF has performed contract research for Biogen MA Inc, and Boehringer Ingelheim. WF has been an invited speaker at Boehringer Ingelheim, Biogen MA Inc, Danone, Eisai, WebMD Neurology (Medscape), Springer Healthcare. WF is consultant to Oxford Health Policy Forum CIC, Roche, and Biogen MA Inc. WF participated in advisory boards of Biogen MA Inc and Roche. All funding is paid to her institution. WF was associate editor of Alzheimer, Research & Therapy in 2020/2021. WF is associate editor at Brain.

CC receives research support from: Biogen, EISAI, Alector and Parabon. The funders of the study had no role in the collection, analysis, or interpretation of data; in the writing of the report; or in the decision to submit the paper for publication. CC is a member of the advisory board of Vivid genetics, Halia Therapeutics and ADx Healthcare.

HZ has served at scientific advisory boards for Denali, Roche Diagnostics, Wave, Samumed, Siemens Healthineers, Pinteon Therapeutics and CogRx, has given lectures in symposia sponsored by Fujirebio, Alzecure and Biogen, and is a co-founder of Brain Biomarker Solutions in Gothenburg AB (BBS), which is a part of the GU Ventures Incubator Program (outside submitted work). KBlennow has served as a consultant, at advisory boards, or at data monitoring committees for Abcam, Axon, Biogen, JOMDD/Shimadzu. Julius Clinical, Lilly, MagQu, Novartis, Roche Diagnostics, and Siemens Healthineers, and is a co-founder of Brain Biomarker Solutions in Gothenburg AB (BBS), which is a part of the GU Ventures Incubator Program.

OAA is a consultant to HealthLytix, and received speaker’s honorarium from Lundbeck and Sunovion. SE has served at scientific advisory boards for Biogen, Danone, Eisai, icometrix, Pfizer, Novartis, Nutricia, Roche and has received unrestricted research grants from ADx Neurosciences and Janssen Pharmaceutica.

CC receives research support from: Biogen, EISAI, Alector, GSK and Parabon. The funders of the study had no role in the collection, analysis, or interpretation of data; in the writing of the report; or in the decision to submit the paper for publication. CC is a member of the advisory board of Vivid genetics, Halia Therapeutics and ADx Healthcare.

HHampel is an employee of Eisai Inc. and serves as Senior Associate Editor for the Journal

Alzheimer’s & Dementia; during the past three years he had received lecture fees from Servier, Biogen and Roche, research grants from Pfizer, Avid, and MSD Avenir (paid to the institution), travel funding from Eisai, Functional Neuromodulation, Axovant, Eli Lilly and company, Takeda and Zinfandel, GE-Healthcare and Oryzon Genomics, consultancy fees from Qynapse, Jung Diagnostics, Cytox Ltd., Axovant, Anavex, Takeda and Zinfandel, GE Healthcare, Oryzon Genomics, and Functional Neuromodulation, and participated in scientific advisory boards of Functional Neuromodulation, Axovant, Eisai, Eli Lilly and company, Cytox Ltd., GE Healthcare, Takeda and Zinfandel, Oryzon Genomics and Roche Diagnostics.

DA participated in advisory boards from Fujirebio-Europe and Roche Diagnostics and received speaker honoraria from Fujirebio-Europe, Roche Diagnostics, Nutricia, Krka Farmacéutica S.L., Zambon S.A.U. and Esteve Pharmaceuticals S.A.

OG reports consulting fees from Eli Lilly, grants to his institution from Actelion, and prescreening activities for Julius Clinical/Toyama.

ABK has been a PI in the drug trials Roche BN29553, Boehringer-Ingelheim 1346.0023 and is PI in Novo Nordisk NN6535-4730.

AL has received personal fees for advisory board services and/or speaker honoraria from Fujirebio-Europe, Roche Diagnostics, Nutricia,Krka Farmacéutica SL, Biogen and Zambon.

PSJ has received personal fees for advisory board from Roche Diagnostics and Zambon.

GS participated in one advisory board meeting from Biogen.

MI is a paid consultant to BioArctic AB. KS is editor at Acta Neuropathologica and associate editor at Alzheimer’s Research & Therapy.

HZ has served at scientific advisory boards for Denali, Roche Diagnostics, Wave, Samumed, Siemens Healthineers, Pinteon Therapeutics and CogRx, has given lectures in symposia sponsored by Fujirebio, Alzecure and Biogen, and is a co-founder of Brain Biomarker Solutions in Gothenburg AB (BBS), which is a part of the GU Ventures Incubator Program (outside submitted work).

All other authors declare no financial interests or potential conflicts of interest.

