## Supplemental text, figures and authors. for "Genome-wide meta-analysis for Alzheimer’s disease cerebrospinal fluid biomarkers"

### Supplementary Information

### Table of content


| | |
|---|---|
| Supplementary Text | |
| Genetic architecture | 2 |
| GWAS results for stratified subgroups | 2 |
| Gene prioritization | 2 |
| Supplementary references | 3 |
| Supplementary figures | 4 |
| Supplementary list of authors | 18 |

### Supplementary results

*Genetic architecture*

To test which of the two proteins has a more comparable genetic background to AD, genetic correlations were calculated with previously published AD GWAS summary statistics, both for Aβ42 and pTau (Supplementary Table 4). Unfortunately, these results were inconclusive, presumably due to the low SNP-heritability for AD diagnosis.

*GWAS rsults for stratified subgroups*

Explorative meta-analyses were repeated stratified for *APOE* (*APOE* ε4 carriers (n=3,240) vs. *APOE* ε4 non-carriers (n=3,201)) and amyloid status (Amyloid normal levels (n=3,182) vs. amyloid abnormal levels (n=3,775)) for stage 1 (QQ plots and lambda shown in Supplementary Figure 9 and 10), of which the results are visualized in Supplementary Figures 11 and 12, and detailed in Supplementary Table 5. *APOE* ε4 carriers still harbored a strong *APOE* signal (Z=-17.92; $p$=8.59x10$^{-72}$), representing the dosage effect of the *APOE* ε4 allele, where rs429358-C decreased Aβ42 protein levels. The 3q28 (*GMNC*) locus was observed for pTau in both the *APOE* ε4 carriers (Z=5.63; $p$=1.85x10$^{-8}$) and non-carriers (Z=7.05; $p$=2.29x10$^{-12}$). We furthermore report a novel locus mapping to chromosome region 7q11.22 for Aβ42 (Z=-5.49; $p$=4.12x10$^{-8}$) in the *APOE* ε4 non-carriers.

Stratified for amyloid status, *APOE* remained strongly associated with decreased CSF Aβ42 levels, both within the abnormal-amyloid-level (Z=-10.03; $p$=1.14x10$^{-23}$) and normal-amyloid-level subgroups (Z=-14.49; $p$=1.40x10$^{-47}$). By contrast, *APOE* increased pTau levels in individuals with normal amyloid levels (Z=-8.53; $p$=1.50x10$^{-17}$), but not in those with abnormal amyloid levels (Z=-2.81; $p$=0.034). The 3q28 (*GMNC*) locus was significantly associated to pTau levels in individuals with both normal and abnormal amyloid levels. A novel locus (12q13.3) was observed for individuals with abnormal amyloid levels, decreasing Aβ42 protein levels (Z=-5.47; $p$=4.50x10$^{-8}$).

*Gene prioritization*

Besides the well-established causal *APOE* gene for the *APOE* locus, the genes *CR1* and *BIN1* have also been indicated as most likely causal genes in previous studies for the AD loci on genomic locations 1q32.2 and 2q14.3, respectively. As this previous work is based on thorough functional experiments, we therefore here only focused with the computational-based tool FUMA [1], on the replicated loci for which the causal gene has not been determined. These loci are the 3q28 and 16q24.2 associations for pTau. We report the most promising causal genes by interpreting positional mapping information, gene-based association tests and eQTL annotations in brain and immune tissue and cell types based on publicly available data (see Methods).

The lead SNP for the 3q28 locus is an intergenic variant 59 kb upstream of *GMNC,* the gene physically closest to the significant association signal. *GMNC* is the only gene within this locus showing an association to pTau based on the aggregated effect of 139 variants ($p$=5.00x$10^{-10}$; Supplementary Table 6). No significant eQTLs were annotated for significant variants within this locus. The lead SNP rs4843559 of 16q24.2 is an intronic variant in *C16orf95*, for which also a significant variant-aggregation association ($p$=4.03x$10^{-9}$) is observed (Supplementary Table 6). Rs4843559 is furthermore a significant eQTL in blood, increasing *ZCCHC14* gene expression.

The variant aggregation analysis showed no newly associated genes, besides the expected associations for genes in the *APOE* locus, *GMNC* and *C16orf95* (Supplementary Table 6).

### Supplementary figures

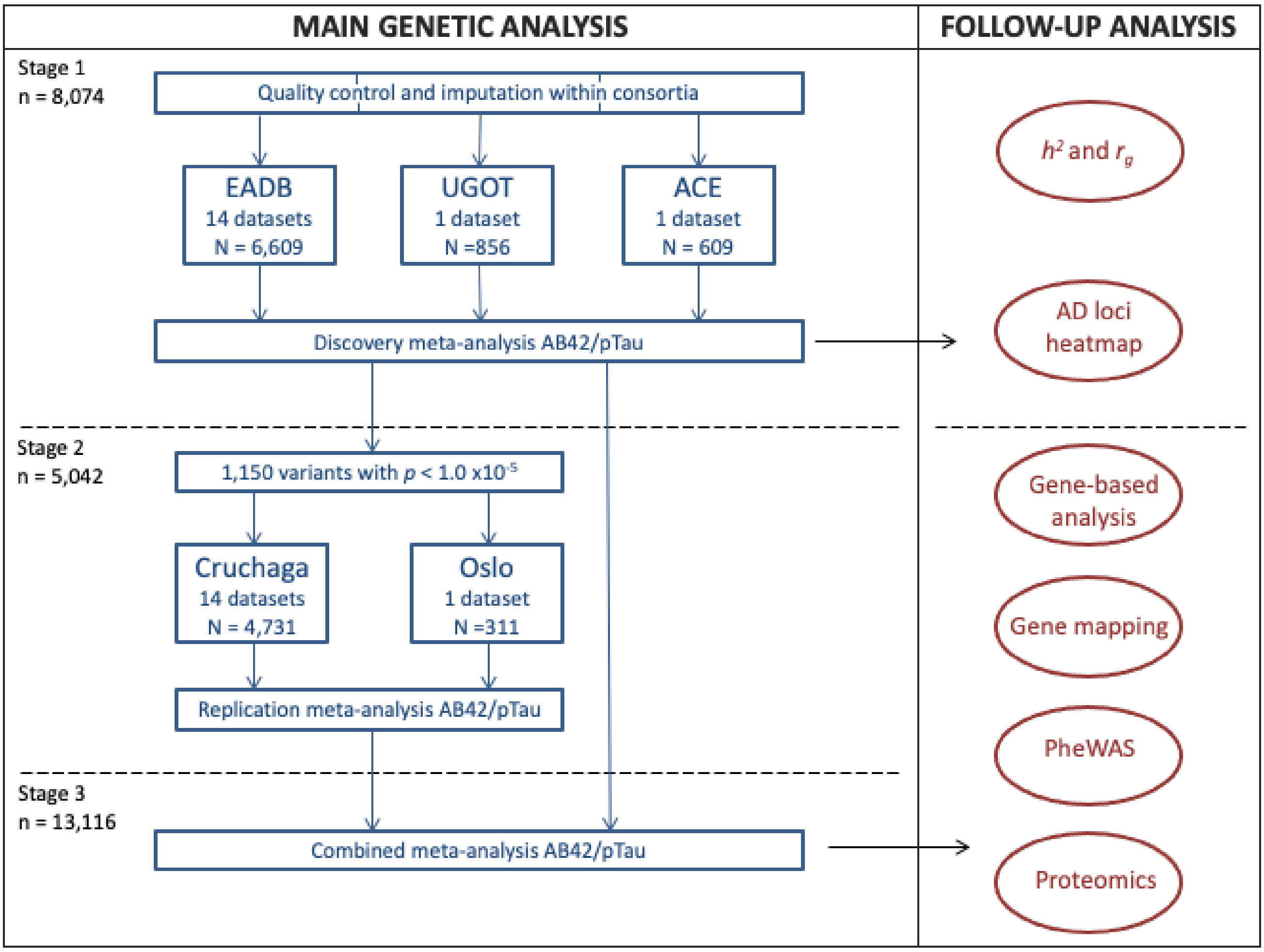


**Figure 1.** Study design of GWAS meta-analysis, and follow up analyses to explore genetic architecture and biological implications. For the follow-up analyses strategies reported in the first two red ovals, it was preferred to maintain n per variant similar in size. For the remaining follow-up analyses the most powerful results were preferred as input.

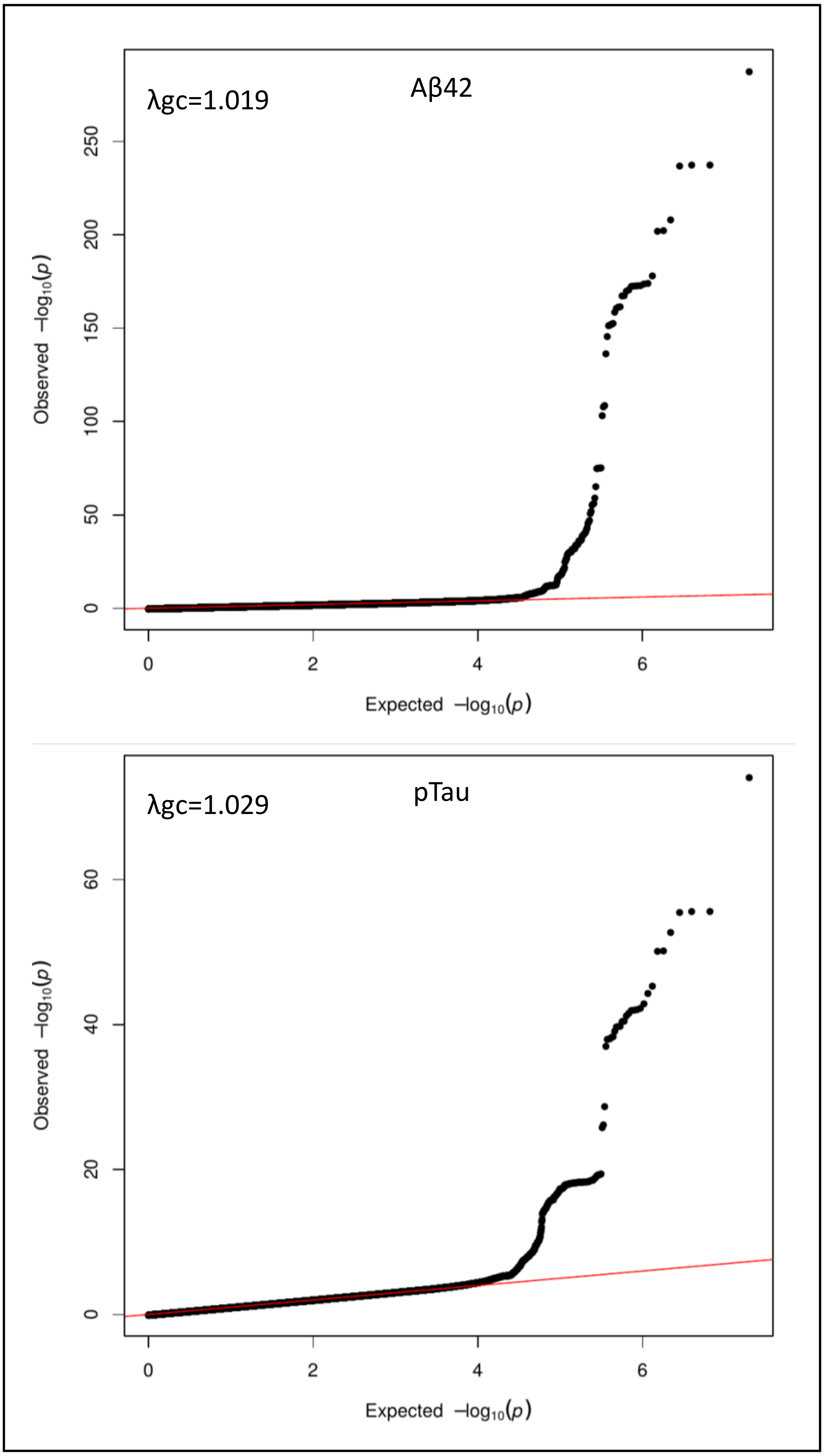


**Figure 2.** QQ plots of Aβ42 and pTau in stage 1. A graphical representation of the deviation of the observed P values from the null hypothesis.

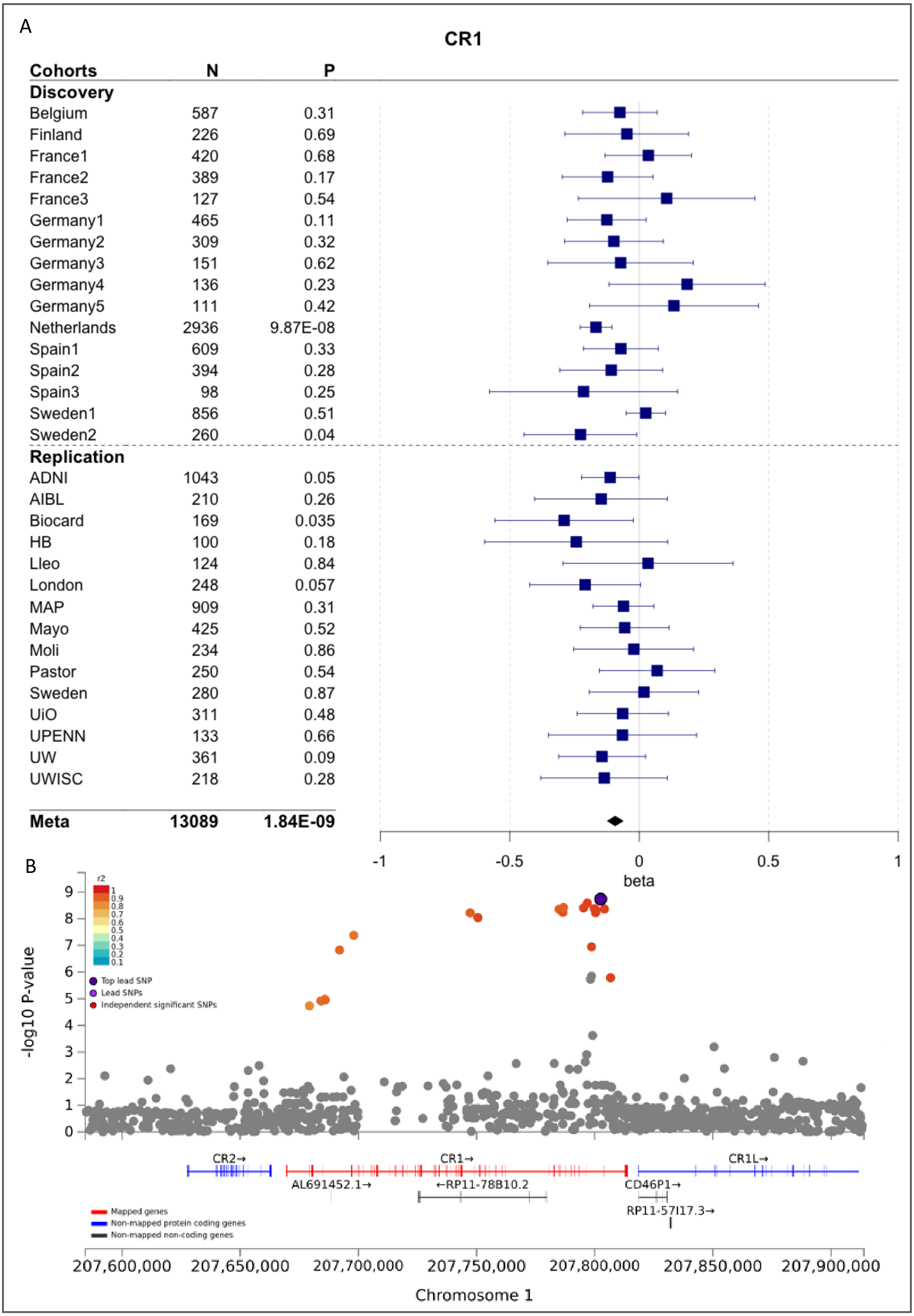


**Figure 3.** Forest (A) and LocusZoom (B) plots of *CR1* locus associating for AB42. Only the variants with an r2 > 0.6 with the lead SNP are color-coded.

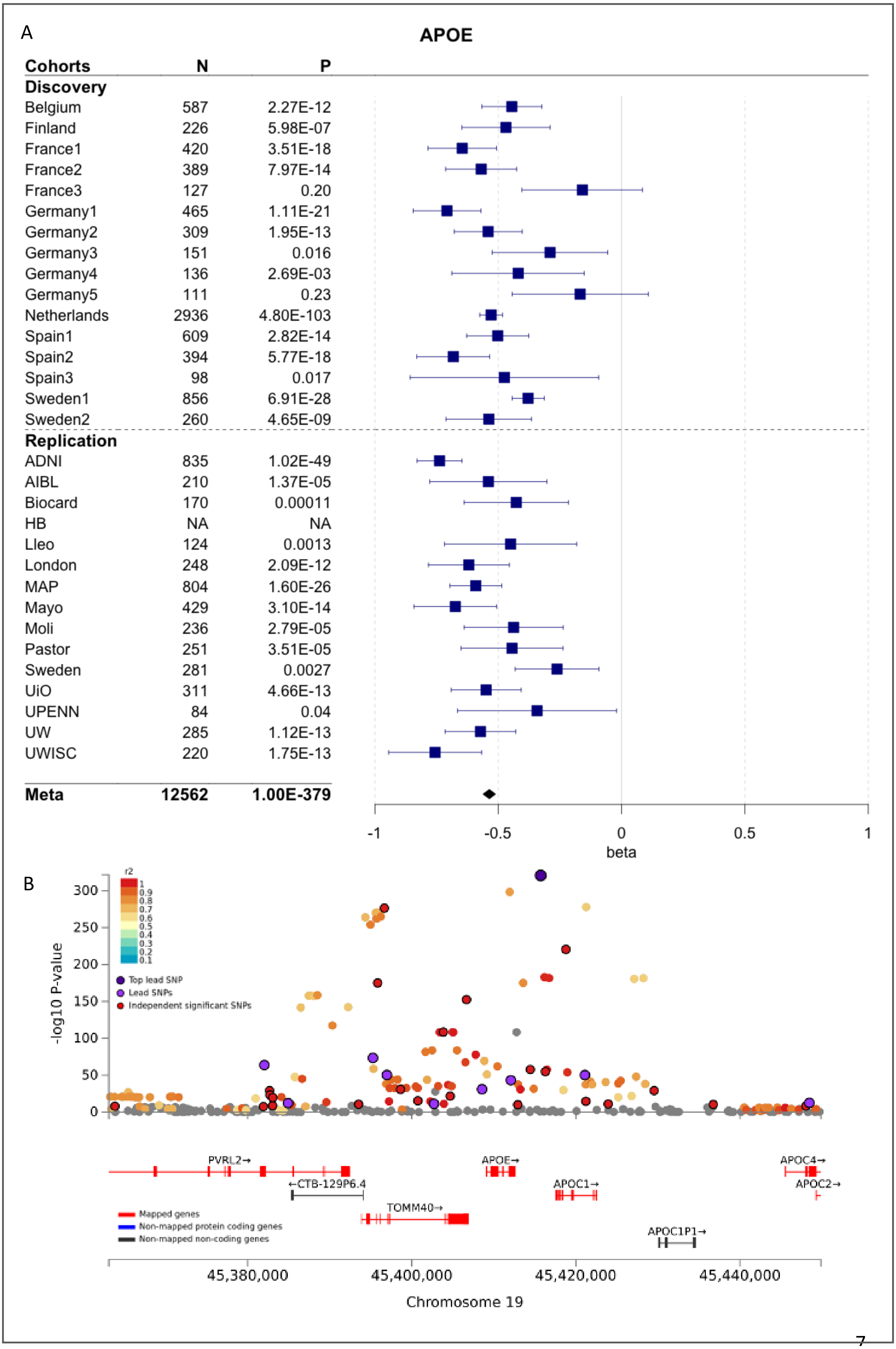


**Figure 4.** Forest (A) and LocusZoom (B) plots of *APOE* locus associating for AB42. Only the variants with an r2 > 0.6 with the lead SNP are color-coded.

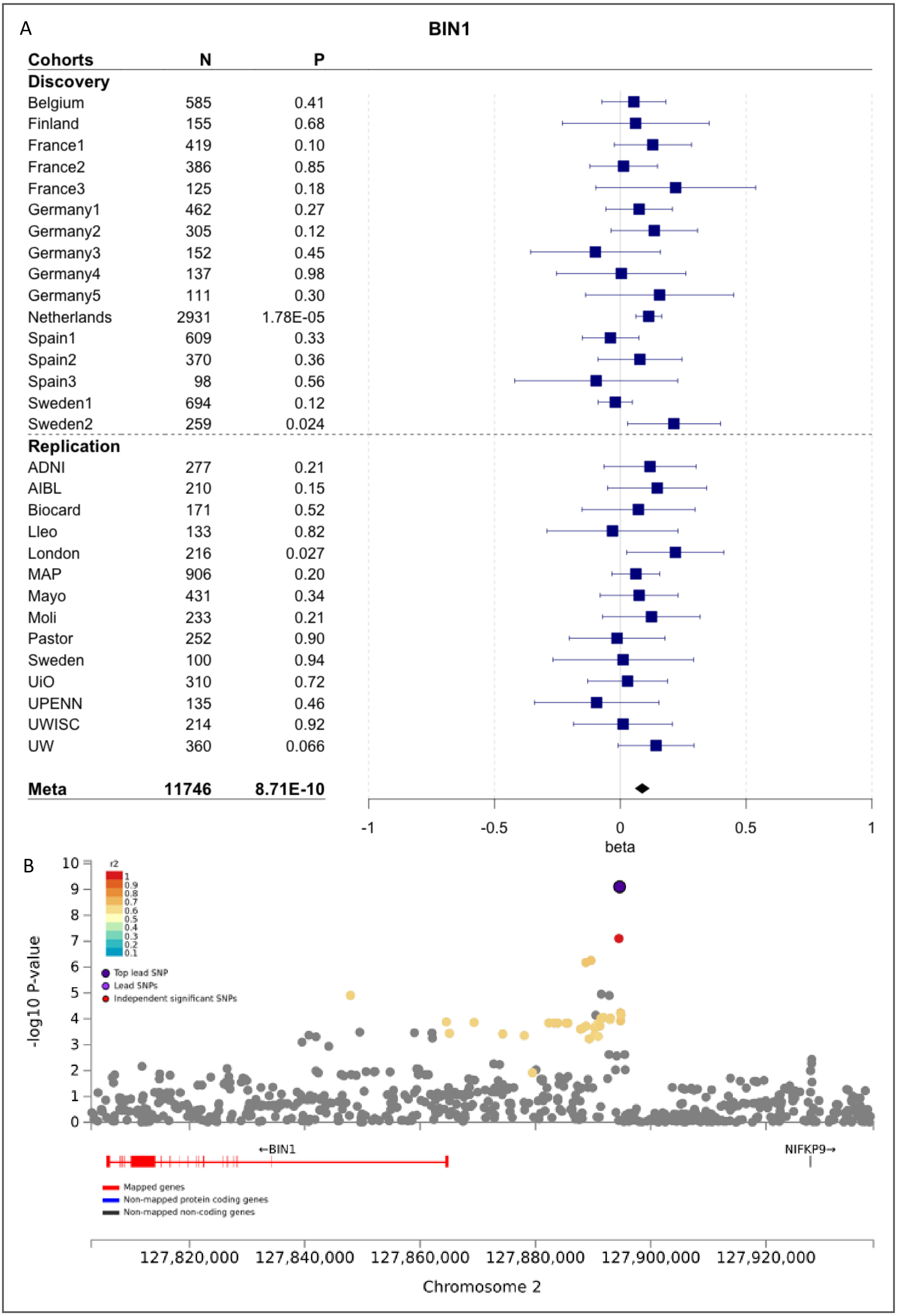


**Figure 5.** Forest (A) and LocusZoom (B) plots of *BIN1* locus associating for pTau. Only the variants with an r2 > 0.6 with the lead SNP are color-coded.

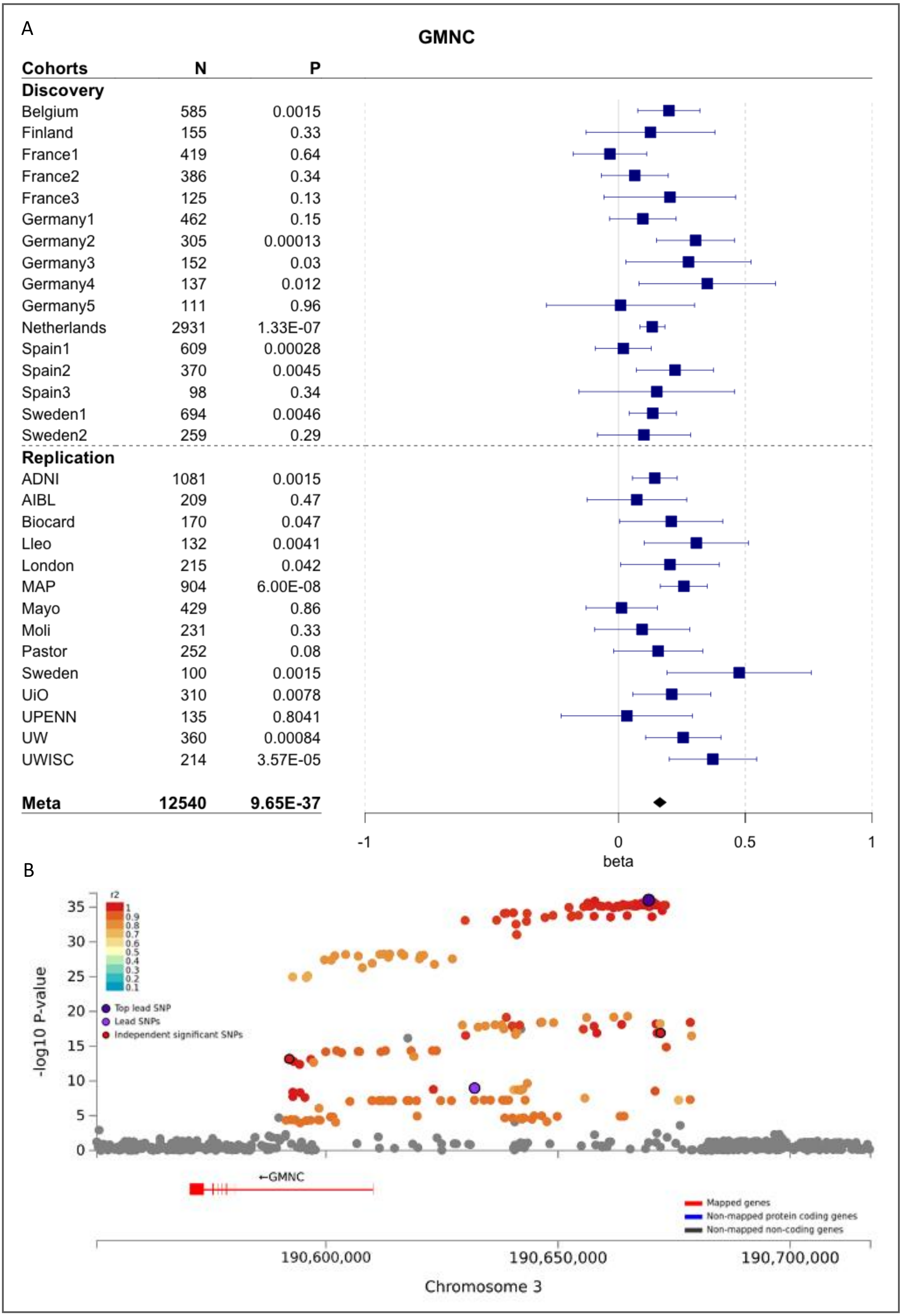


**Figure 6.** Forest (A) and LocusZoom (B) plots of *GMNC* locus associating for pTau. Only the variants with an r2 > 0.6 with the lead SNP are color-coded. The significance level for variants in high LD can vary due to differences in N as a part of the variants were not available in the replication cohorts.

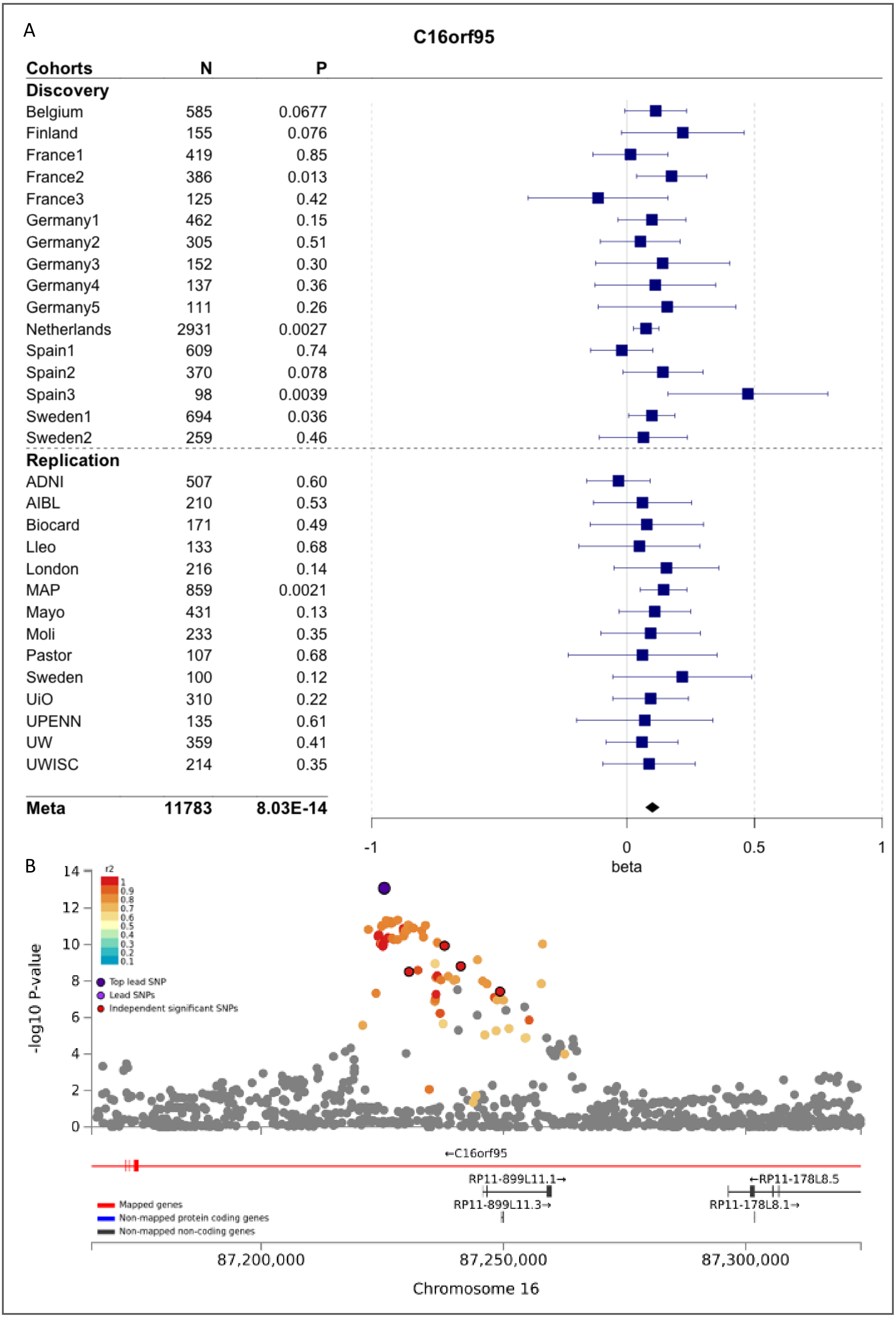


**Figure 7.** Forest (A) and LocusZoom (B) plots of *C16orf95* locus associating for pTau. Only the variants with an r2 > 0.6 with the lead SNP are color-coded.

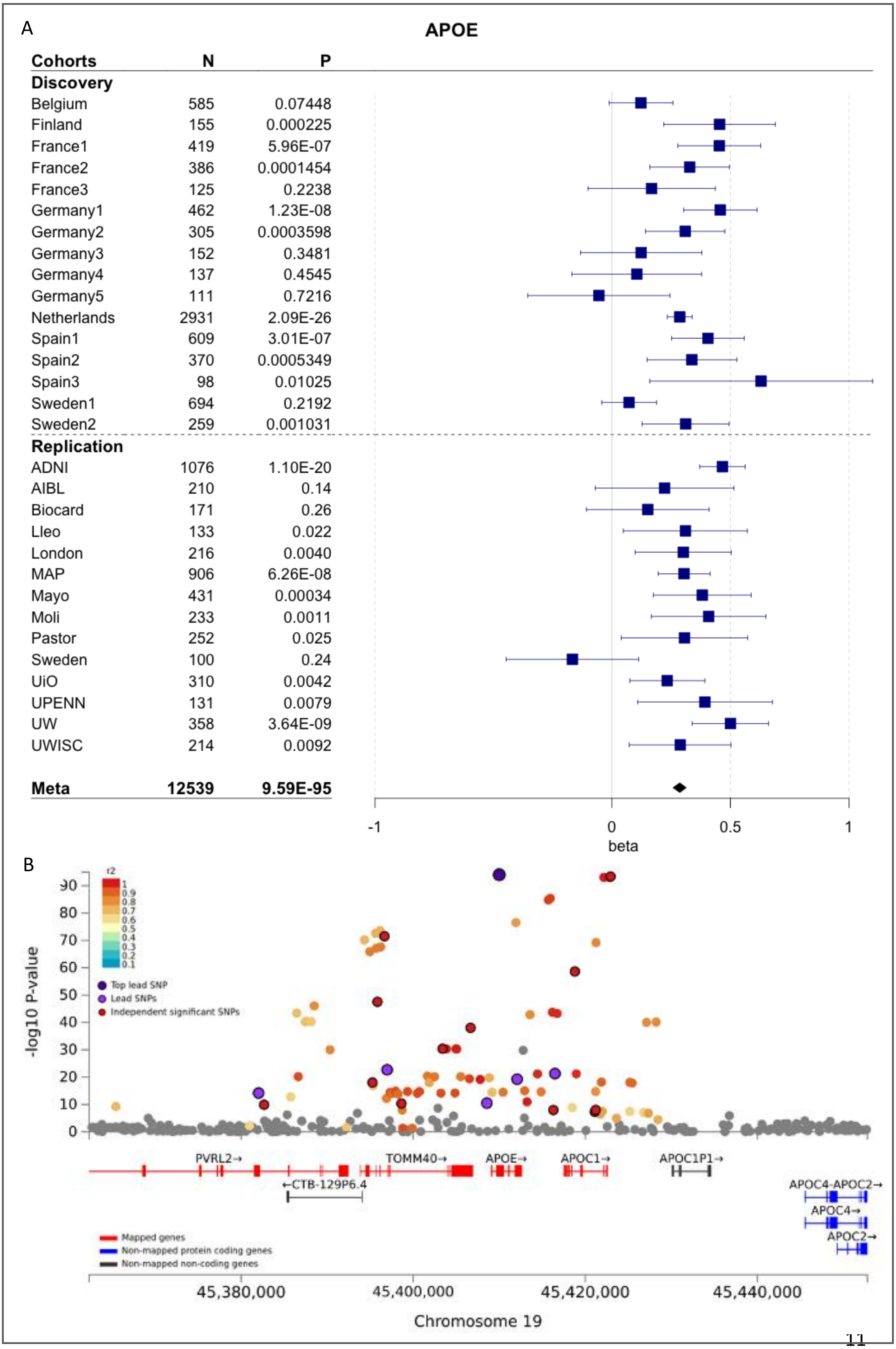




**Figure 8**. Forest (A) and LocusZoom (B) plots of *APOE* locus associating for pTau. Only the variants with an r2 > 0.6 with the lead SNP are color-coded.

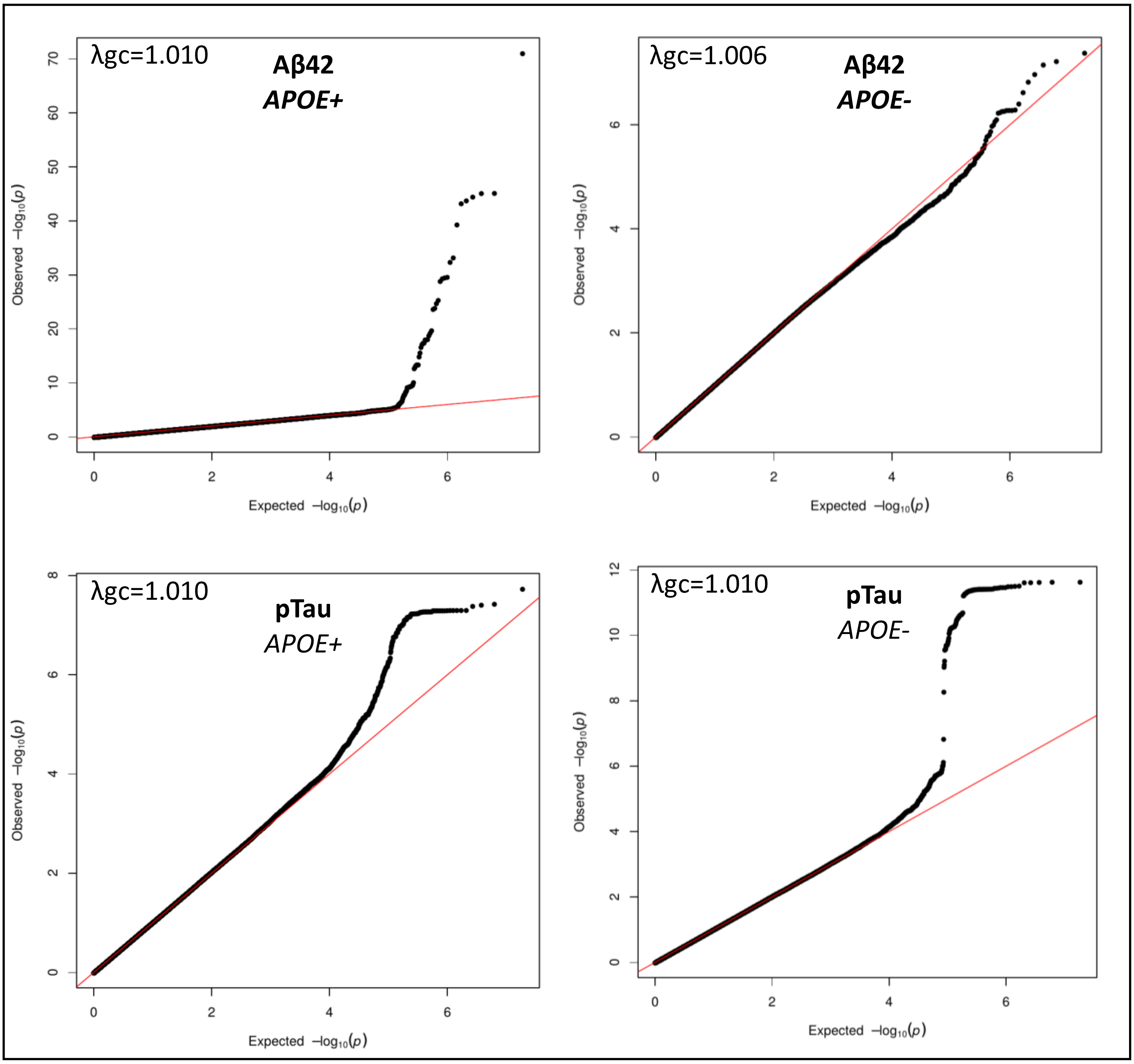


**Figure 9.** QQ plots of Aβ42 and pTau stratified for APOE4 status. A graphical representation of the deviation of the observed P values from the null hypothesis.

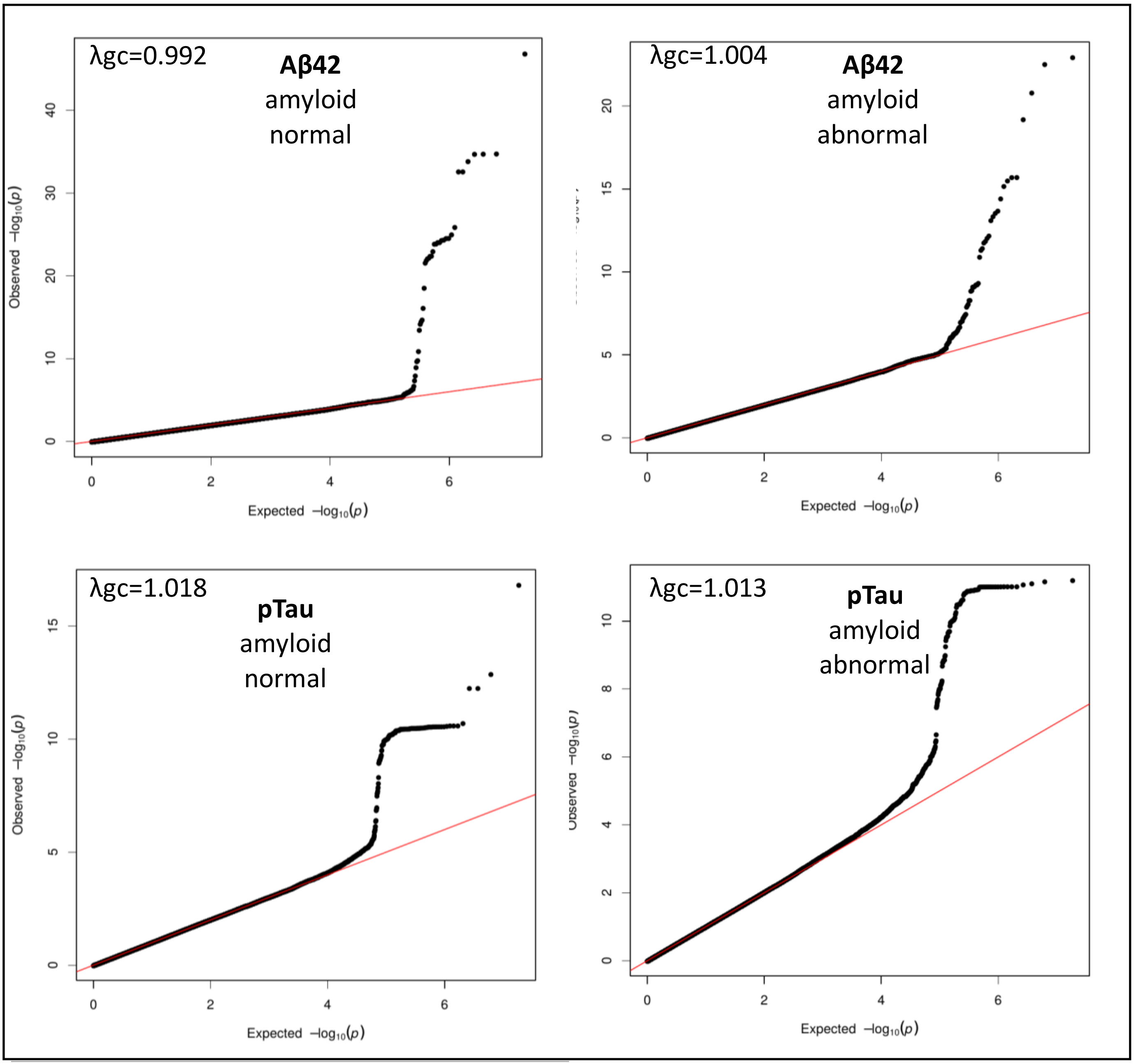


**Figure 10.** QQ plots of Aβ42 and pTau stratified for amyloid status. A graphical representation of the deviation of the observed P values from the null hypothesis.

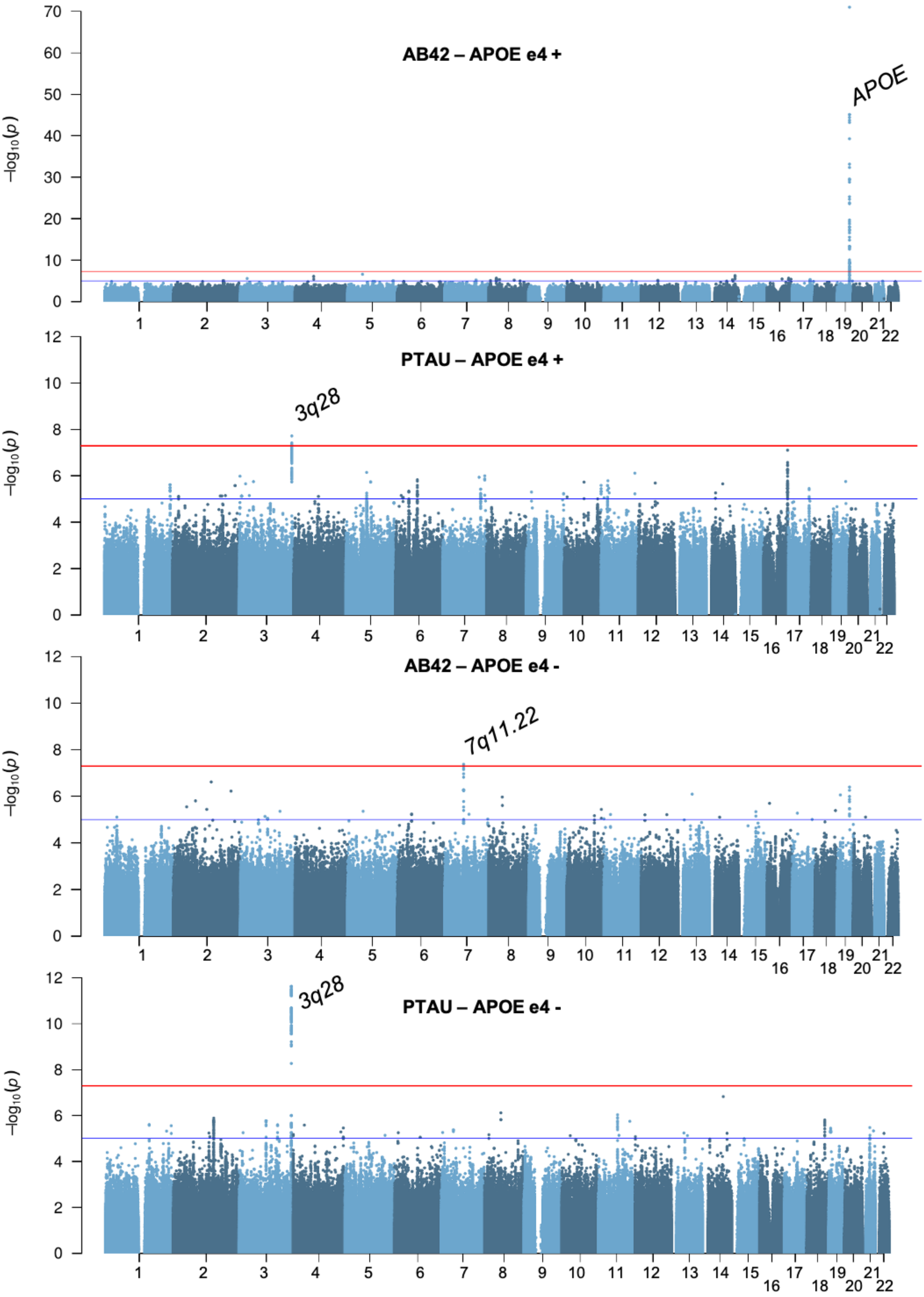


**Figure 11.** Manhattan plots of association results for stratification based on APOE e4 status.

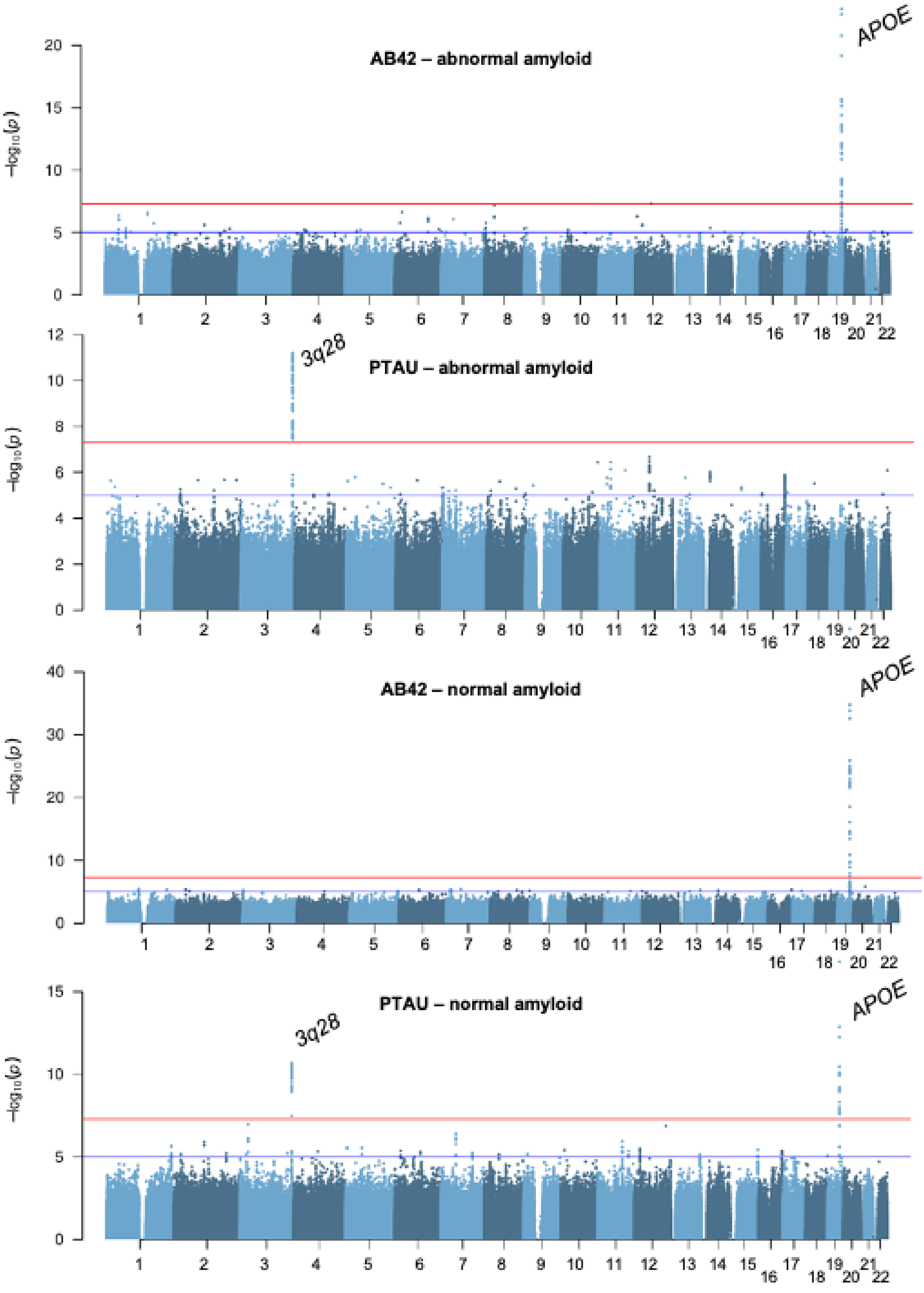


**Figure 12.** Manhattan plots of association results for stratification based on amyloid status in CSF.

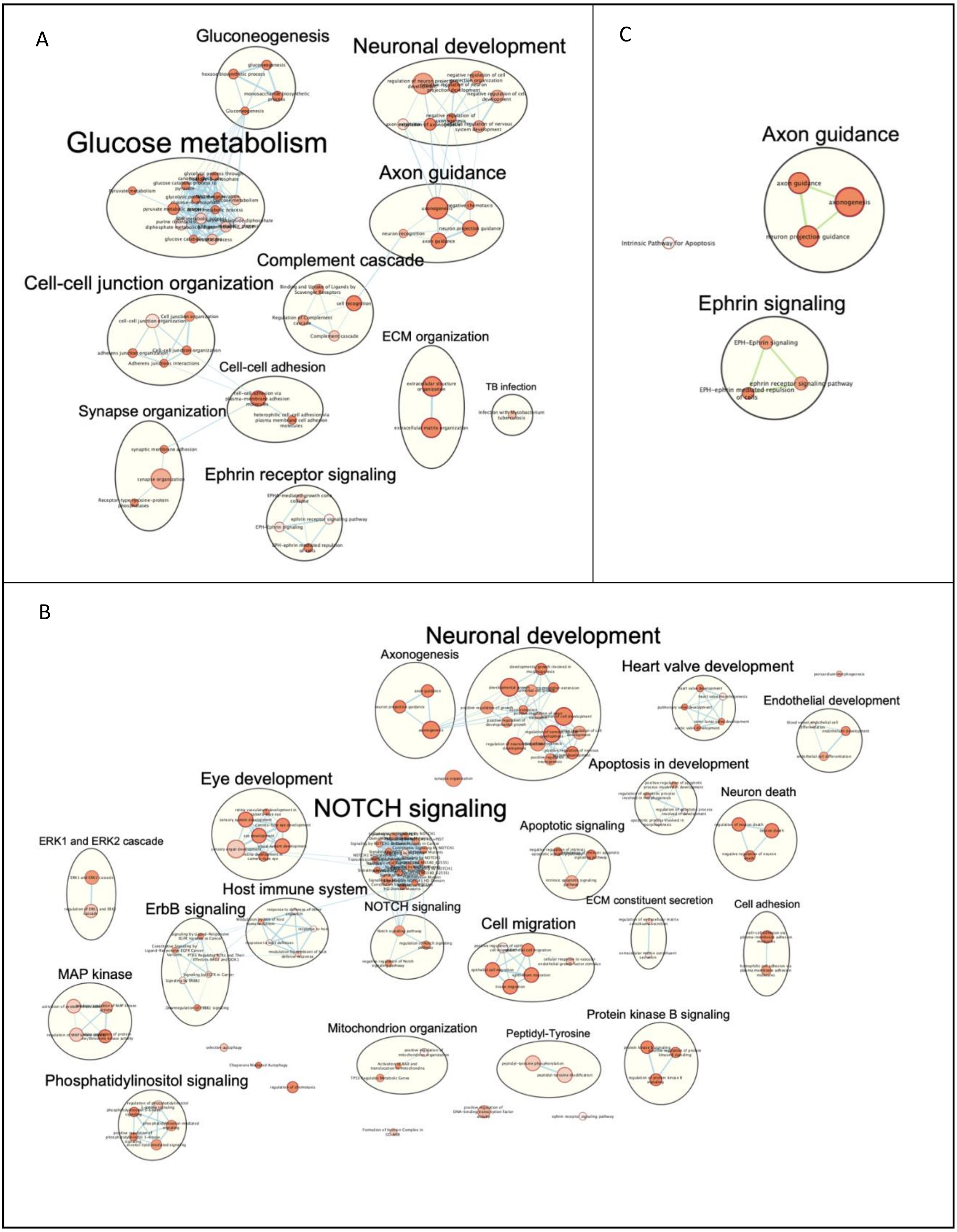


**Figure 13.** Pathway analyses based on CSF-protein analyses of *GMNC* variants within the EMIF dataset (A), the WUSTL dataset (B). (C) displays the overlapping functional pathways between the different datasets.

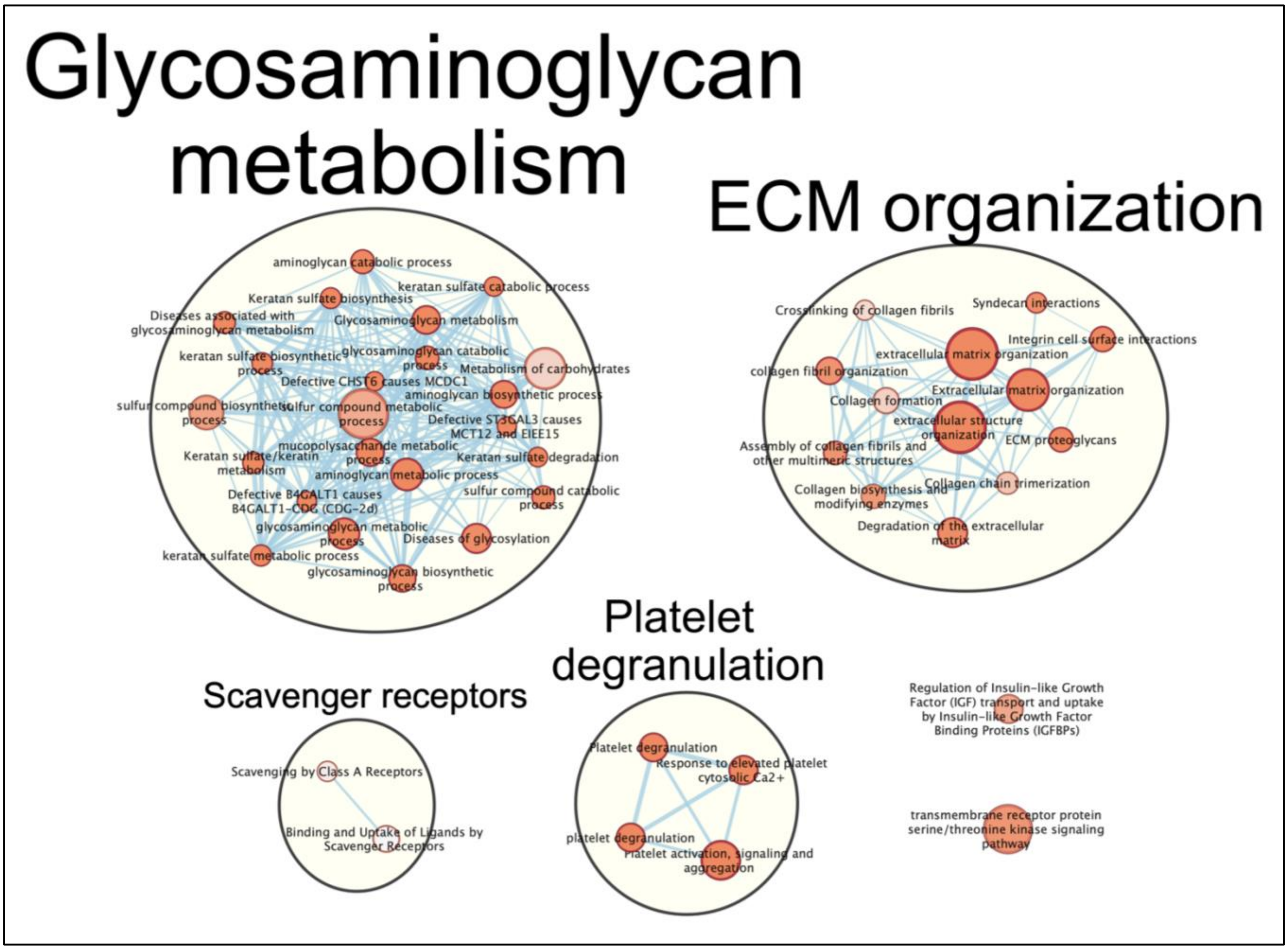


**Figure 14.** *Pathway analyses based on CSF-protein analyses of GMNC variants within the EMIF dataset.*

### Supplementary list of authors


#### EADB

Altenstein Slawek[1,2], Brosseron Frederic[3,4], Burow Lena[5], Cetindag Arda[6], Ewers Michael[7,8], Fenski Friederike[6], Fliessbach Klaus[3,4], Glanz Wenzel[9], Janowitz Daniel[8], Kilimann Ingo[10,11], Maier Franziska[12], Metzger Coraline D.[9,13], Munk Matthias H.[14,15], Preis Lukas[6], Spruth Eike J[1,6].

1. German Center for Neurodegenerative Diseases (DZNE), Berlin, Germany.
2. Department of Psychiatry and Psychotherapy, Charité, Berlin, Germany.
3. German Center for Neurodegenerative Diseases (DZNE), Bonn, Germany.
4. University of Bonn Medical Center, Dept. of Neurodegenerative Disease and Geriatric Psychiatry/Psychiatry, Bonn, Germany.
5. Department of Psychiatry and Psychotherapy, University Hospital, LMU Munich, Munich, Germany.
6. Charité-Universitätsmedizin Berlin, Campus Benjamin Franklin, Department of Psychiatry, Berlin, Germany.
7. German Center for Neurodegenerative Diseases (DZNE, Munich), Munich, Germany.
8. Institute for Stroke and Dementia Research (ISD), University Hospital, LMU Munich, Munich, Germany.
9. German Center for Neurodegenerative Diseases (DZNE), Magdeburg, Germany.
10. German Center for Neurodegenerative Diseases (DZNE), Rostock, Germany.
11. Department of Psychosomatic Medicine, Rostock University Medical Center, Rostock, Germany.
12. Department of Psychiatry, University of Cologne, Medical Faculty, Cologne, Germany.
13. Institute of Cognitive Neurology and Dementia Research (IKND), Otto-von-Guericke University, Magdeburg, Germany.
14. Institute of Cognitive Neurology and Dementia Research (IKND), Otto-von-Guericke University, Magdeburg, Germany.
15. Section for Dementia Research, Hertie Institute for Clinical Brain Research and Department of Psychiatry and Psychotherapy, University of Tübingen, Tübingen, Germany.

#### The GR@ACE study group

Aguilar Miquel[1,2], Aguilera Nuria[3,] Alarcon Emilio[3], Alegret Montserrat[3,4], Boada Mercè[3,4], Buendia Mar[3], Cano Amanda[3], Cañabate Pilar[3,4], Carracedo Angel[6,7], Corbatón-Anchuelo A[8], de Rojas Itziar[3,4], Diego Susana[3], Espinosa Ana[3,4], Gailhajenet Anna[3], García-González Pablo[3,4], Guitart Marina[3], González-Pérez Antonio[9], Ibarria Marta[3], Lafuente Asunción[3], Macias Juan[10], Maroñas Olalla[6], Martín Elvira[3], Martínez Maria Teresa[8], Marquié Marta[3,4], Montrreal Laura[3], Moreno-Grau Sonia[3,4], Moreno Mariona[3], Raúl Nuñez-Llaves[3], Olivé Clàudia[1] , Orellana Adelina[3,4], Ortega Gemma[3,4], Pancho Ana[3], Pelejà Ester[3], Pérez-Cordon Alba[3], Pineda Juan A[10], Puerta Raquel[3], Preckler Silvia[3], Quintela Inés[5], Real Luis Miguel[5,10], Rosende-Roca Maitee[3], Ruiz Agustín[3,4], Sáez Maria Eugenia[9], Sanabria Angela[3,4], Serrano-Rios Manuel[8], Sotolongo-Grau Oscar[3], Tárraga Luís[3,4], Valero Sergi[3,4], Vargas Liliana[1].

1. Memory Disorders Unit, Department of Neurology, Hospital Universitari Mutua de Terrassa, Terrassa, Spain.
2. Fundació per a la Recerca Biomèdica i Social Mútua de Terrassa, Terrassa, Spain.
3. Research Center and Memory clinic. ACE Alzheimer Center Barcelona, Universitat Internacional de Catalunya, Spain.
4. CIBERNED, Center for Networked Biomedical Research on Neurodegenerative Diseases, National Institute of Health Carlos III, Ministry of Economy and Competitiveness, Spain,
5. Dep. of Surgery, Biochemistry and Molecular Biology, School of Medicine. University of Málaga. Málaga, Spain,
6. Grupo de Medicina Xenómica, Centro Nacional de Genotipado (CEGEN-PRB5-ISCIII). Universidad de Santiago de Compostela, Santiago de Compostela, Spain.
7. Fundación Pública Galega de Medicina Xenómica- CIBERER-IDIS, Santiago de Compostela, Spain.
8. Centro de Investigación Biomédica en Red de Diabetes y Enfermedades Metabólicas Asociadas, CIBERDEM, Spain, Hospital Clínico San Carlos, Madrid, Spain,
9. CAEBI. Centro Andaluz de Estudios Bioinformáticos, Sevilla, Spain
10. Unidad Clínica de Enfermedades Infecciosas y Microbiología. Hospital Universitario de Valme, Sevilla, Spain.
